# Disparities in SARS-CoV-2 case rates by ethnicity, religion, measures of socio-economic position, English proficiency, and self-reported disability: cohort study of 39 million people in England during the Alpha and Delta waves

**DOI:** 10.1101/2022.03.02.22271762

**Authors:** Tim Larsen, Matthew L. Bosworth, Daniel Ayoubkhani, Ryan Schofield, Raghib Ali, Kamlesh Khunti, Ann Sarah Walker, Myer Glickman, Vahé Nafilyan

## Abstract

**Objective:** To examine socio-demographic disparities in SARS-CoV-2 case rates during the second (Alpha) and third (Delta) waves of the COVID-19 pandemic.

**Design:** Retrospective, population-based cohort study.

**Setting:** Resident population of England.

**Participants:** 39,006,194 people aged 10 years and over who were enumerated at the 2011 Census, registered with the National Health Service (NHS) and alive on 1 September 2020.

**Main outcome measures:** Testing positive for SARS-CoV-2 during the second wave (1 September 2020 to 22 May 2021) or third wave (23 May to 10 December 2021) of the pandemic. We calculated age-standardised case rates by socio-demographic characteristics and used logistic regression models to estimate adjusted odds ratios (ORs).

**Results:** During the study period, 5,767,584 individuals tested positive for SARS-CoV-2. In the second wave, the fully-adjusted odds of having a positive test, relative to the White British group, were highest for the Bangladeshi (OR: 1.88, 95% CI 1.86 to 1.90) and Pakistani (1.81, 1.79 to 1.82) ethnic groups. Relative to the Christian group, Muslim and Sikh religious groups had fully-adjusted ORs of 1.58 (1.57 to 1.59) and 1.74 (1.72 to 1.76), respectively. Greater area deprivation, disadvantaged socio-economic position, living in a care home and low English language proficiency were also associated with higher odds of having a positive test. However, the disparities between groups varied over time. Being Christian, White British, non-disabled, and from a more advantaged socio-economic position were all associated with increased odds of testing positive during the third wave.

**Conclusion:** There are large socio-demographic disparities on SARS-CoV-2 cases which have varied between different waves of the pandemic. Research is now urgently needed to understand why these disparities exist to inform policy interventions in future waves or pandemics.

**What is already known on this topic:** People with pre-existing health conditions or disability, ethnic minority groups, the elderly, some religious groups, people with low socio-economic status, and those living in deprived areas have been disproportionately affected by the COVID-19 pandemic in terms of risk of infection and adverse outcomes.

**What this study adds:** Using linked data on 39 million people in England, we found that during the second wave, COVID-19 case rates were highest among the Bangladeshi and Pakistani ethnic groups, the Muslim religious group, individuals from deprived areas and of low socio-economic position; during the third wave, being Christian, White British, non-disabled, and from a more advantaged socio-economic position were all associated with increased odds of receiving a positive test

Adjusting for geographical factors, socio-demographic characteristics, and pre-pandemic health status explained some, but not all, of the excess risk

When stratifying the dataset by broad age groups, the odds of receiving a positive test remained higher among the Bangladeshi and Pakistani ethnic groups aged 65 years and over during the third wave, which may partly explain the continued elevated mortality rates in these groups

## Introduction

As of 18 February 2022, there have been over 418 million confirmed cases of SARS-CoV-2 globally, with more than 160,000 deaths in the United Kingdom [1] [2]. Whilst the COVID-19 pandemic has affected all areas of the UK, some groups have been disproportionally affected. Rates of COVID-19 related hospitalisation and death have been higher among the elderly, people with pre-existing health conditions or disability [3] [4] [5], ethnic minority groups [6] [7] [8], some religious groups [9], people with low socio-economic status [10], and those living in care homes [11], large households [12], and deprived areas [13] [14] [15].

Less is known about socio-demographic disparities in infection rates. Research using data from the Coronavirus Infection Survey, a large household survey representative of the UK community population, has demonstrated that several factors were associated with SARS-CoV-2 positivity during the second wave and early part of the third wave in the UK [16] [17] [18]. Other studies have also highlighted non-white ethnicity, male sex and living in an urban or more deprived area as risk factors for testing positive [6] [19] [20]. However, large-scale studies using national population-level data sources that adjust for key confounding variables to understand the drivers of increased infection rates are lacking [21]. Given that socio-demographic disparities in severe COVID-19 outcomes appear to be largely driven by differences in infection rates, there is a clear evidence gap with which to inform national policies to reduce infection risk.

In this study, we examined differences in case rates across a range of socio-demographic characteristics, including ethnicity, religion, measures of socio-economic position and self-reported disability status using population-level administrative and Census data.

## Methods

### Study data

We linked national SARS-CoV-2 test results obtained via Pillar 1 (swab testing in UK Health Security Agency laboratories and NHS hospitals for those with a clinical need, and health and care workers) and Pillar 2 (swab testing for the wider population, as set out in government guidance) to the Office for National Statistics (ONS) Public Health Data Asset (PHDA) using NHS number. The ONS PHDA is a linked data resource combining the 2011 Census, death registrations, General Practice Extraction Service (GPES) Data for Pandemic Planning and Research (GDPPR) [22] and Hospital Episode Statistics (HES) [23]. To obtain NHS numbers, we linked the 2011 Census to the 2011-2013 NHS Patient Registers using deterministic and probabilistic matching, with an overall linkage rate of 94.6%. The NHS numbers in national testing data were incomplete, with missing values for 21% of records. To retrieve additional NHS numbers, we linked the testing data to the NHS Personal Demographics Service (PDS) using deterministic matching, achieving a linkage rate of 91.4%.

The study population consisted of all people aged ≥10 years living in England, who were enumerated at the 2011 Census, registered with a general practitioner (GP) surgery in November 2019, and alive on 1 September 2020. The cohort comprised to 39,006,194, 78.4% of the mid-year 2020 population estimate of people aged ≥10 years in England [24].

We used national testing data up to 10 December 2021. Out of all test results, 83.0% were linked to the ONS PHDA. It was not possible to calculate case rates and odds ratios for the first wave as mass testing was not available.

### Exposures and covariates

All individual-level socio-demographic characteristics (sex, age, ethnic group, religious affiliation, disability status, educational attainment, National Statistics Socio-economic Classification (NS-SEC) of the household reference person, English language proficiency, country of birth) were available from the 2011 Census. Place of residence (region within England and Rural-Urban Classification [25]) and area-based deprivation (English Indices of Deprivation, 2019 [26]) were derived based on postcodes held in GP records. Care home residence was retrieved from the 2019 NHS Patient Register. Pre-existing health conditions were derived from GDPPR data as in the QCOVID risk prediction model [3]. We included the number of pre-existing conditions and separate adjustments for learning disability, as it could directly affect the exposure to SARS-CoV-2 [27]. We also adjusted for body mass index, as a categorical variable with a category for missing values. All variables included in the analyses are listed in Table S1 in the supplementary material.

### Outcome

The outcome was receiving a positive test result (polymerase chain reaction [PCR] or lateral flow device [LFD], including LFD positives that were not confirmed by PCR) for SARS-CoV-2 in a dataset of one row per individual within the study population. We excluded any positive tests that occurred within 120 days of an initial positive test from the same individual as these may have been part of the same infection episode [28]. We classified tests up to and including 22 May 2021 as having occurred in the second wave of the COVID-19 pandemic, with tests from 23 May 2021 to 10 December 2021 classified as being in the third wave [17].

### Statistical analyses

We estimated age-standardised SARS-CoV-2 case rates as the number positive cases per 100,000 person-weeks at risk, stratified by socio-demographic characteristics, standardised to the 2013 European Standard Population [29]. Rates were calculated weekly and for the second (1 September 2020 to 22 May 2021) and third (23 May to 10 December 2021) waves of the pandemic.

For each exposure, we compared odds ratios (ORs) for testing positive for SARS-CoV-2 estimated from logistic regression models adjusted in a stepwise manner for different sets of covariates. We estimated ORs adjusted for: sex and age (model 1); sex, age and geographical variables (region and Rural-Urban Classification) (model 2); and sex, age, geography, socio-demographic characteristics (ethnicity, Indices of Deprivation, educational attainment, household tenure, and care-home residence status), and self-reported disability status and the number of pre-existing health conditions (model 3). Due to the significant overlap between ethnicity and religion, we did not include religion in the covariate set. The number of pre-existing health conditions was included as a proxy for contact with the healthcare system, which may affect the risk of SARS-CoV-2 infection or lead to shielding. Contact with the healthcare system would also make the individual more likely to be tested for SARS-CoV-2.

We explored how differences in the risk of testing positive for SARS-CoV-2 changed over the course of the pandemic by fitting separate models for wave two and wave three. We also fitted separate models for those aged under 65 years and those 65 years and over.

All analyses were conducted using R version 3.5.

### Patient and public involvement

No patients involved.

## Results

Out of the 39,006,194 individuals in our study population, 52.1% were female, the mean age was 47.6 (SD: 21.1) years, 81.7% identified as White British, 4.8% as White Other, 2.7% as Indian, 59.5% as Christian, 25.5% as having no religious affiliation, and 5% as Muslim (Table S1). Between 1 September 2020 and 10 December 2021, 5,767,584 people (14.8% of the study population) living in England aged ≥10 years had tested positive for SARS-CoV-2, of which 46,484 (0.8%; 0.1% of the total study population) had an infection episode in both the second and third waves of the pandemic

During the second wave, the largest differences in rates of testing positive for SARS-CoV-2 were observed for ethnicity; rates were highest in the Bangladeshi and Pakistani ethnic groups at 382.4 and 373.8 per 100,000 person-weeks, respectively, and in the Chinese ethnic group, with 90.8 cases per 100,000 person-weeks. During the third wave, however, the White British ethnic group had the highest rate at 359.7 cases per 100,000 person-weeks (Table 2). There were also notable disparities in case rates by religious affiliation. During the second wave of the pandemic, case rates per 100,000 person-weeks were highest for people who identified as Muslim (334.9) or Sikh (321.6). Rates were lowest for people in the ‘Other Religion’ group (142.9) and the Buddhist group (143.3). During the third wave, those who identified as Christian had the highest rates at 353.8 cases per 100,000 person-weeks, whereas the lowest rates were found in the Buddhist and Muslim groups at 221.4 and 226.7 cases per 100,000 person-weeks, respectively.

**Table 1:**
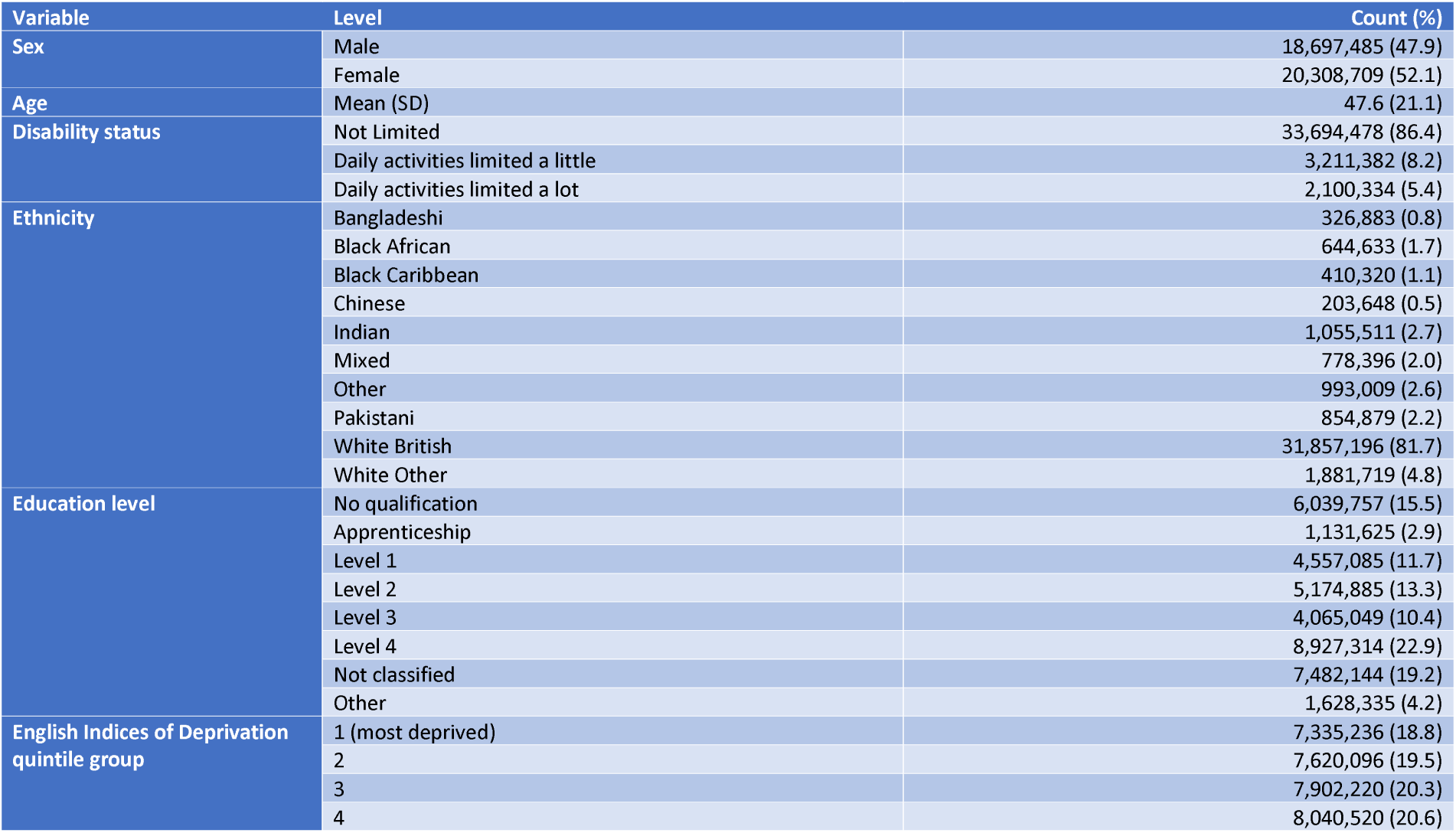
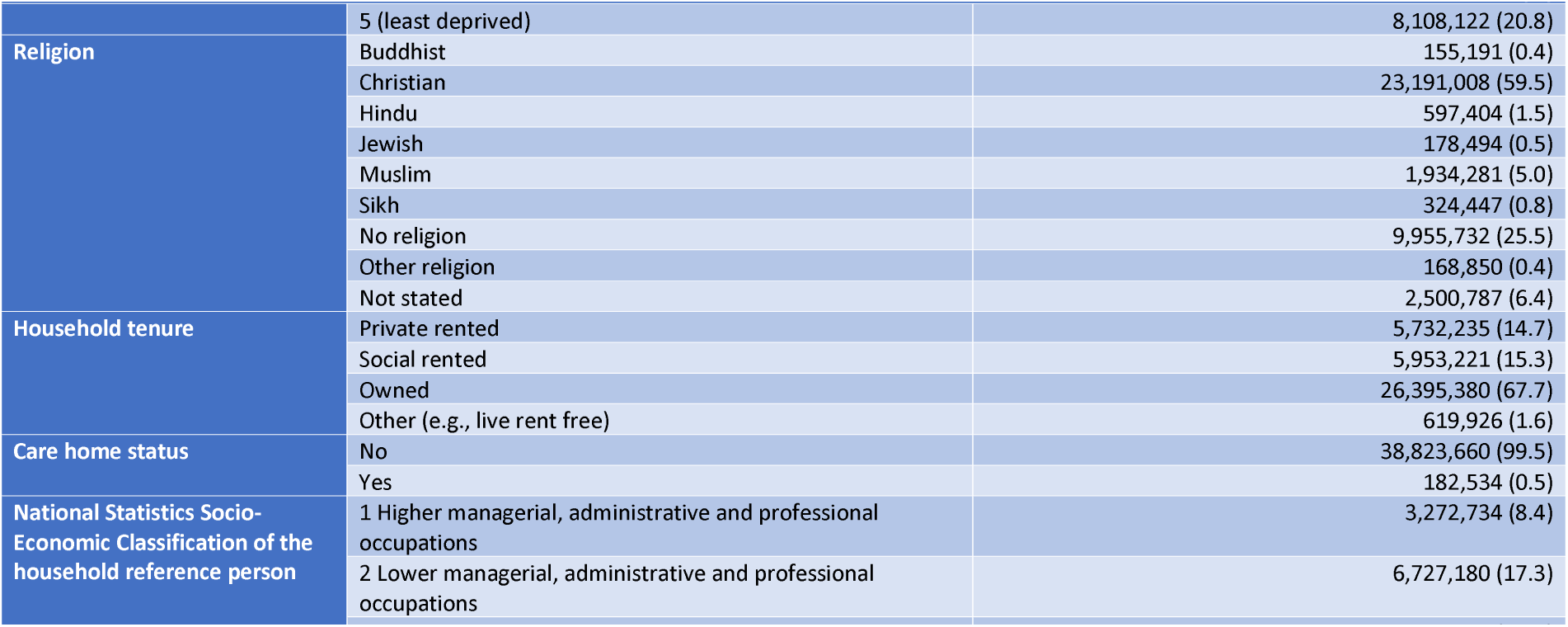
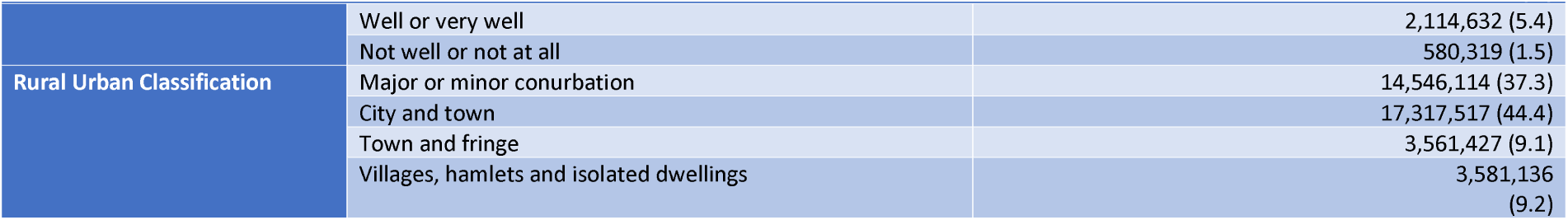
Characteristics of the study population

**Table 2:** Age-standardised SARS-CoV-2 case rates (per 100,000 person-weeks) by socio-demographic characteristics and wave of the pandemic

|  |  | Wave two (1 September 2020 to 22 May 2021) |  |  |  | Wave three (23 May 2021 onwards) |  |  |  |
| --- | --- | --- | --- | --- | --- | --- | --- | --- | --- |
| Exposure | Level | Number of cases | Rate | Lower CI | Upper CI | Number of cases | Rate | Lower CI | Upper CI |
| Ethnic group | Bangladeshi | 43,449 | 382.4 | 377.9 | 386.9 | 23,756 | 229.9 | 226.3 | 233.5 |
|  | Black African | 47,855 | 200.2 | 198.0 | 202.4 | 41,958 | 198.4 | 196.2 | 200.5 |
|  | Black Caribbean | 27,748 | 184.6 | 182.3 | 186.8 | 28,941 | 266.4 | 263.2 | 269.5 |
|  | Chinese | 6,811 | 90.8 | 88.5 | 93.0 | 9,031 | 162.5 | 159.0 | 165.9 |
|  | Indian | 102,001 | 267.3 | 265.6 | 269.0 | 80,550 | 265.9 | 264.0 | 267.8 |
|  | Mixed | 55,724 | 183.5 | 181.5 | 185.5 | 88,670 | 303.5 | 301.0 | 305.9 |
|  | Other | 87,798 | 238.0 | 236.3 | 239.7 | 68,648 | 225.5 | 223.7 | 227.3 |
|  | Pakistani | 110,638 | 373.8 | 371.2 | 376.4 | 62,132 | 233.1 | 231.0 | 235.2 |
|  | White British | 1,851,398 | 165.3 | 165.0 | 165.5 | 2,824,792 | 359.7 | 359.2 | 360.1 |
|  | White Other | 115,184 | 166.7 | 165.7 | 167.8 | 136,984 | 260.3 | 258.8 | 261.8 |
| Religious group | Buddhist | 8,043 | 143.3 | 139.9 | 146.7 | 8,860 | 221.4 | 216.3 | 226.4 |
|  | Christian | 1,406,889 | 177.3 | 177.0 | 177.6 | 1,920,206 | 353.8 | 353.3 | 354.3 |
|  | Hindu | 49,248 | 227.3 | 225.2 | 229.4 | 45,158 | 265.2 | 262.7 | 267.7 |
|  | Jewish | 11,730 | 189.8 | 186.3 | 193.3 | 13,298 | 293.1 | 288.0 | 298.1 |
|  | Muslim | 228,476 | 334.9 | 333.3 | 336.5 | 139,064 | 226.7 | 225.4 | 228.1 |
|  | Sikh | 37,471 | 321.6 | 318.3 | 325.0 | 26,322 | 286.0 | 282.5 | 289.5 |
|  | No religion | 564,183 | 147.2 | 146.8 | 147.6 | 1,000,330 | 336.2 | 335.6 | 336.9 |
|  | Other religion | 8,284 | 142.9 | 139.4 | 146.3 | 10,725 | 267.6 | 261.6 | 273.7 |
|  | Not stated | 134,282 | 151.9 | 151.1 | 152.7 | 201,499 | 304.9 | 303.5 | 306.2 |
| Country of | UK | 2,089,141 | 171.7 | 171.5 | 171.9 | 3,081,174 | 345.0 | 344.6 | 345.4 |

| Exposure | Level | Wave two (1 September 2020 to 22 May 2021) |  |  |  | Wave three (23 May 2021 onwards) |  |  |  |
| --- | --- | --- | --- | --- | --- | --- | --- | --- | --- |
|  |  | Number of cases | Rate | Lower CI | Upper CI | Number of cases | Rate | Lower CI | Upper CI |
| birth | Non-UK | 359,465 | 203.1 | 202.4 | 203.9 | 284,288 | 238.2 | 237.1 | 239.4 |
| English Indices of Deprivation quintile group | 1 (most deprived) | 581,068 | 218.7 | 218.1 | 219.2 | 644,804 | 311.8 | 311.0 | 312.6 |
|  | 2 | 527,010 | 191.2 | 190.7 | 191.7 | 649,484 | 317.3 | 316.6 | 318.1 |
|  | 3 | 472,664 | 168.5 | 168.0 | 169.0 | 666,824 | 331.5 | 330.7 | 332.3 |
|  | 4 | 450,659 | 160.3 | 159.8 | 160.8 | 690,861 | 347.5 | 346.7 | 348.3 |
|  | 5 (least deprived) | 417,205 | 148.0 | 147.6 | 148.5 | 713,489 | 358.1 | 357.3 | 359.0 |
| National Statistics Socio-Economic Classification of the household reference person | Higher managerial, administrative and professional occupations | 139,872 | 102.0 | 100.7 | 103.3 | 229,509 | 211.3 | 208.8 | 213.7 |
|  | Lower managerial, administrative and professional occupations | 369,283 | 128.3 | 127.7 | 128.9 | 498,925 | 228.8 | 227.7 | 229.9 |
|  | Intermediate occupations | 250,852 | 141.0 | 140.3 | 141.6 | 299,393 | 228.8 | 227.8 | 229.8 |
|  | Small employers and own account workers | 153,985 | 127.6 | 126.7 | 128.6 | 163,628 | 189.7 | 188.1 | 191.3 |
|  | Lower supervisory and technical occupations | 134,166 | 142.5 | 141.7 | 143.3 | 144,680 | 213.1 | 211.8 | 214.3 |
|  | Semi-routine occupations | 299,012 | 158.0 | 157.4 | 158.6 | 293,050 | 211.9 | 211.1 | 212.7 |
|  | Routine occupations | 208,805 | 145.7 | 145.1 | 146.4 | 203,723 | 202.9 | 202.0 | 203.9 |
|  | Never worked and long-term unemployed | 110,573 | 153.8 | 152.9 | 154.8 | 92,397 | 158.8 | 157.8 | 159.8 |
| Education level | No qualification | 356,433 | 150.4 | 149.8 | 151.0 | 258,959 | 175.6 | 174.8 | 176.4 |
|  | Apprenticeship | 58,991 | 138.0 | 136.7 | 139.3 | 64,967 | 224.2 | 222.1 | 226.2 |
|  | Level 1 | 300,887 | 144.3 | 143.7 | 144.9 | 335,274 | 211.2 | 210.5 | 211.9 |
|  | Level 2 | 347,997 | 142.4 | 141.9 | 142.9 | 410,814 | 219.1 | 218.4 | 219.8 |
|  | Level 3 | 270,548 | 137.7 | 137.2 | 138.3 | 343,028 | 223.3 | 222.5 | 224.1 |
|  |  | Number of cases | Rate | Lower CI | Upper CI | Number of cases | Rate | Lower CI | Upper CI |
|  | Level 4 | 456,144 | 119.3 | 118.6 | 119.9 | 656,680 | 212.4 | 211.4 | 213.5 |
|  | Other | 108,109 | 156.4 | 155.4 | 157.5 | 83,920 | 165.7 | 164.3 | 167.1 |
| Rural-Urban Classification | Cities and towns | 1,005,795 | 162.3 | 162.0 | 162.6 | 1,565,237 | 350.0 | 349.4 | 350.5 |
|  | Major or minor conurbations | 1,129,050 | 215.0 | 214.6 | 215.3 | 1,223,891 | 308.7 | 308.1 | 309.2 |
|  | Towns and fringes | 171,629 | 139.9 | 139.2 | 140.5 | 307,205 | 360.1 | 358.8 | 361.4 |
|  | Villages, hamlets and isolated dwellings | 142,132 | 118.8 | 118.1 | 119.4 | 269,129 | 332.3 | 331.0 | 333.7 |
| Disability status | Non-disabled - not limited | 2,147,056 | 174.0 | 173.8 | 174.2 | 3,134,229 | 337.6 | 337.3 | 338.0 |
|  | Disabled - limited a little | 173,719 | 162.9 | 161.9 | 163.9 | 146,457 | 272.0 | 270.1 | 273.9 |
|  | Disabled - limited a lot | 127,831 | 159.9 | 158.7 | 161.1 | 84,776 | 212.6 | 210.6 | 214.6 |
| Region | North East | 132,204 | 194.7 | 193.6 | 195.7 | 204,171 | 420.4 | 418.6 | 422.2 |
|  | North West | 400,613 | 218.2 | 217.5 | 218.8 | 487,936 | 363.6 | 362.5 | 364.6 |
|  | Yorkshire and the Humber | 270,098 | 192.0 | 191.3 | 192.7 | 384,690 | 375.8 | 374.6 | 377.0 |
|  | East Midlands | 218,633 | 177.9 | 177.1 | 178.6 | 320,396 | 363.1 | 361.8 | 364.4 |
|  | West Midlands | 282,221 | 193.6 | 192.9 | 194.3 | 354,840 | 332.9 | 331.8 | 334.0 |
|  | East of England | 250,486 | 158.4 | 157.8 | 159.1 | 352,650 | 308.6 | 307.6 | 309.7 |
|  | London | 406,794 | 205.4 | 204.7 | 206.0 | 371,473 | 241.6 | 240.8 | 242.4 |
|  | South East | 343,314 | 150.6 | 150.1 | 151.1 | 525,119 | 319.4 | 318.6 | 320.3 |
|  | South West | 144,243 | 101.3 | 100.8 | 101.9 | 364,187 | 370.2 | 368.9 | 371.4 |
| English language proficiency | Main language | 2,211,286 | 171.6 | 171.4 | 171.8 | 3,191,368 | 342.2 | 341.9 | 342.6 |
|  | Speak English very well or well | 187,607 | 239.4 | 238.2 | 240.6 | 140,685 | 228.2 | 226.8 | 229.5 |
|  |  | Number of cases | Rate | Lower CI | Upper CI | Number of cases | Rate | Lower CI | Upper CI |
|  | Do not speak English well or at all | 49,713 | 238.8 | 236.2 | 241.5 | 33,409 | 194.4 | 191.6 | 197.2 |
| Household tenure | Private rented | 367,298 | 168.4 | 167.8 | 169.1 | 548,583 | 299.4 | 298.5 | 300.3 |
|  | Social rented | 419,093 | 193.1 | 192.5 | 193.7 | 508,011 | 296.3 | 295.5 | 297.1 |
|  | Owned | 1,601,009 | 177.8 | 177.5 | 178.1 | 2,237,339 | 355.1 | 354.6 | 355.6 |
|  | Other | 38,571 | 169.3 | 167.6 | 171.0 | 50,985 | 300.6 | 298.0 | 303.3 |

CI, confidence interval (95%).

In the second wave, the Bangladeshi ethnic group had the highest odds ratio (OR) of testing positive for SARS-CoV-2 relative to the White British ethnic group (Table 3); adjusting for age and sex only, the OR was 2.19 (95% confidence interval 2.17 to 2.22), whereas the fully-adjusted OR was 1.88 (1.86 to 1.90). Geography, socio-demographic factors and pre-pandemic health status accounted for 26.1% of the elevated odds of testing positive for SARS-CoV-2 among the Bangladeshi ethnic group during the second wave of the pandemic. During the third wave, however, the odds of testing positive for SARS-CoV-2 were lower for all ethnic minority groups compared with the White British group.

**Table 3:** Adjusted odds ratios of receiving a positive test for SARS-CoV-2 by sociodemographic characteristics and wave of the pandemic

| Exposure | Group | Wave two (1 September 2020 to 22 May 2021) |  |  | Wave three (23 May 2021 onwards) |  |  |
| --- | --- | --- | --- | --- | --- | --- | --- |
|  |  | OR (Model 1) | OR (Model 2) | OR (Model 3) | OR (Model 1) | OR (Model 2) | OR (Model 3) |
| Sex | Female | 1 (ref) | 1 (ref) | 1 (ref) | 1 (ref) | 1 (ref) | 1 (ref) |
|  | Male | 0.86 [0.86 - 0.86] | 0.86 [0.86 - 0.86] | 0.87 [0.87 - 0.87] | 0.91 [0.91 - 0.91] | 0.91 [0.91 - 0.91] | 0.91 [0.91 - 0.91] |
| Age group | 10-19 | 0.83 [0.82 - 0.83] | 0.84 [0.84 - 0.85] | 0.84 [0.84 - 0.85] | 2.20 [2.19 - 2.21] | 2.20 [2.19 - 2.21] | 2.20 [2.19 - 2.21] |
|  | 20-29 | 1.19 [1.18 - 1.19] | 1.20 [1.20 - 1.21] | 1.20 [1.20 - 1.21] | 1.19 [1.18 - 1.19] | 1.18 [1.17 - 1.18] | 1.18 [1.17 - 1.18] |
|  | 30-39 | 1 (ref) | 1 (ref) | 1 (ref) | 1 (ref) | 1 (ref) | 1 (ref) |
|  | 40-49 | 0.96 [0.96 - 0.97] | 0.98 [0.97 - 0.98] | 0.98 [0.97 - 0.98] | 1.11 [1.11 - 1.12] | 1.12 [1.11 - 1.12] | 1.12 [1.11 - 1.12] |
|  | 50-59 | 0.87 [0.86 - 0.87] | 0.90 [0.89 - 0.90] | 0.90 [0.89 - 0.90] | 0.71 [0.70 - 0.71] | 0.70 [0.70 - 0.70] | 0.70 [0.70 - 0.70] |
|  | 60-69 | 0.56 [0.55 - 0.56] | 0.58 [0.58 - 0.58] | 0.58 [0.58 - 0.58] | 0.43 [0.42 - 0.43] | 0.42 [0.42 - 0.42] | 0.42 [0.42 - 0.42] |
|  | 70-79 | 0.37 [0.37 - 0.37] | 0.39 [0.39 - 0.39] | 0.39 [0.39 - 0.39] | 0.25 [0.25 - 0.26] | 0.25 [0.25 - 0.25] | 0.25 [0.25 - 0.25] |
|  | 80-89 | 0.60 [0.59 - 0.60] | 0.63 [0.63 - 0.64] | 0.63 [0.63 - 0.64] | 0.18 [0.18 - 0.19] | 0.18 [0.18 - 0.18] | 0.18 [0.18 - 0.18] |
|  | 90+ | 1.20 [1.18 - 1.21] | 1.28 [1.27 - 1.29] | 1.28 [1.27 - 1.29] | 0.21 [0.20 - 0.21] | 0.21 [0.20 - 0.21] | 0.21 [0.20 - 0.21] |
| Disability | Not Limited | 1 (ref) | 1 (ref) | 1 (ref) | 1 (ref) | 1 (ref) | 1 (ref) |
|  | Limited a little | 1.02 [1.02 - 1.03] | 1.00 [1.00 - 1.01] | 0.92 [0.91 - 0.92] | 0.84 [0.84 - 0.84] | 0.83 [0.83 - 0.84] | 0.85 [0.85 - 0.86] |
|  | Limited a lot | 1.15 [1.15 - 1.16] | 1.10 [1.09 - 1.11] | 0.93 [0.92 - 0.93] | 0.72 [0.71 - 0.72] | 0.71 [0.70 - 0.71] | 0.76 [0.75 - 0.76] |
| Ethnicity | White British | 1 (ref) | 1 (ref) | 1 (ref) | 1 (ref) | 1 (ref) | 1 (ref) |
|  | Bangladeshi | 2.19 [2.17 - 2.22] | 1.96 [1.94 - 1.98] | 1.88 [1.86 - 1.90] | 0.55 [0.54 - 0.56] | 0.61 [0.60 - 0.62] | 0.64 [0.63 - 0.65] |
|  | Black African | 1.16 [1.15 - 1.17] | 1.06 [1.05 - 1.07] | 1.05 [1.04 - 1.06] | 0.51 [0.50 - 0.51] | 0.57 [0.57 - 0.58] | 0.61 [0.60 - 0.62] |
|  | Black Caribbean | 1.12 [1.10 - 1.13] | 1.01 [1.00 - 1.02] | 0.97 [0.96 - 0.98] | 0.75 [0.74 - 0.76] | 0.86 [0.85 - 0.87] | 0.89 [0.88 - 0.91] |
|  | Chinese | 0.53 [0.51 - 0.54] | 0.49 [0.48 - 0.50] | 0.54 [0.52 - 0.55] | 0.41 [0.41 - 0.42] | 0.44 [0.43 - 0.45] | 0.45 [0.44 - 0.46] |
|  | Indian | 1.65 [1.64 - 1.66] | 1.52 [1.50 - 1.53] | 1.55 [1.54 - 1.57] | 0.72 [0.71 - 0.72] | 0.77 [0.77 - 0.78] | 0.77 [0.76 - 0.77] |
|  | Mixed | 1.11 [1.10 - 1.12] | 1.05 [1.04 - 1.06] | 1.05 [1.04 - 1.06] | 0.83 [0.82 - 0.84] | 0.88 [0.88 - 0.89] | 0.91 [0.90 - 0.92] |

|  |  | Wave two (1 September 2020 to 22 May 2021) |  |  | Wave three (23 May 2021 onwards) |  |  |
| --- | --- | --- | --- | --- | --- | --- | --- |
|  | Other | 1.45 [1.44 - 1.46] | 1.34 [1.33 - 1.35] | 1.35 [1.34 - 1.36] | 0.59 [0.59 - 0.59] | 0.66 [0.65 - 0.66] | 0.69 [0.68 - 0.69] |
|  | Pakistani | 2.17 [2.16 - 2.19] | 1.89 [1.87 - 1.90] | 1.81 [1.79 - 1.82] | 0.57 [0.56 - 0.57] | 0.57 [0.57 - 0.58] | 0.58 [0.58 - 0.59] |
|  | White Other | 1.00 [1.00 - 1.01] | 0.97 [0.96 - 0.97] | 1.00 [1.00 - 1.01] | 0.71 [0.70 - 0.71] | 0.77 [0.76 - 0.77] | 0.81 [0.80 - 0.81] |
| Education level | No qualification | 1 (ref) | 1 (ref) | 1 (ref) | 1 (ref) | 1 (ref) | 1 (ref) |
|  | Apprenticeship | 0.93 [0.92 - 0.94] | 0.98 [0.97 - 0.99] | 1.04 [1.03 - 1.05] | 1.31 [1.30 - 1.33] | 1.29 [1.28 - 1.30] | 1.19 [1.18 - 1.20] |
|  | Level 1 | 0.91 [0.90 - 0.91] | 0.95 [0.94 - 0.95] | 0.98 [0.98 - 0.99] | 1.17 [1.16 - 1.17] | 1.17 [1.16 - 1.17] | 1.11 [1.10 - 1.11] |
|  | Level 2 | 0.90 [0.89 - 0.90] | 0.94 [0.94 - 0.95] | 0.99 [0.99 - 1.00] | 1.21 [1.20 - 1.21] | 1.20 [1.20 - 1.21] | 1.13 [1.12 - 1.13] |
|  | Level 3 | 0.87 [0.86 - 0.87] | 0.92 [0.91 - 0.92] | 0.97 [0.97 - 0.98] | 1.24 [1.23 - 1.24] | 1.24 [1.23 - 1.24] | 1.16 [1.15 - 1.16] |
|  | Level 4 | 0.71 [0.71 - 0.71] | 0.75 [0.74 - 0.75] | 0.79 [0.78 - 0.79] | 1.19 [1.18 - 1.19] | 1.23 [1.23 - 1.24] | 1.17 [1.16 - 1.18] |
|  | Other | 1.02 [1.02 - 1.03] | 1.02 [1.01 - 1.03] | 1.01 [1.00 - 1.02] | 0.92 [0.91 - 0.93] | 0.98 [0.97 - 0.99] | 1.07 [1.06 - 1.08] |
| English Indices of Deprivation quintile group | 1 | 1.49 [1.48 - 1.50] | 1.29 [1.29 - 1.30] | 1.19 [1.19 - 1.20] | 0.86 [0.86 - 0.87] | 0.82 [0.81 - 0.82] | 0.86 [0.86 - 0.86] |
|  | 2 | 1.31 [1.30 - 1.32] | 1.22 [1.22 - 1.23] | 1.16 [1.15 - 1.16] | 0.88 [0.88 - 0.89] | 0.90 [0.89 - 0.90] | 0.92 [0.92 - 0.93] |
|  | 3 | 1.15 [1.14 - 1.15] | 1.14 [1.14 - 1.15] | 1.11 [1.11 - 1.12] | 0.92 [0.92 - 0.93] | 0.94 [0.93 - 0.94] | 0.95 [0.95 - 0.95] |
|  | 4 | 1.09 [1.08 - 1.09] | 1.09 [1.09 - 1.10] | 1.08 [1.07 - 1.08] | 0.97 [0.96 - 0.97] | 0.97 [0.97 - 0.97] | 0.97 [0.97 - 0.98] |
|  | 5 | 1 (ref) | 1 (ref) | 1 (ref) | 1 (ref) | 1 (ref) | 1 (ref) |
| Religion | Christian | 1 (ref) | 1 (ref) | 1 (ref) | 1 (ref) | 1 (ref) | 1 (ref) |
|  | Buddhist | 0.80 [0.78 - 0.81] | 0.79 [0.77 - 0.81] | 0.83 [0.81 - 0.85] | 0.60 [0.59 - 0.62] | 0.65 [0.64 - 0.67] | 0.68 [0.67 - 0.70] |
|  | Hindu | 1.30 [1.29 - 1.31] | 1.22 [1.21 - 1.24] | 1.27 [1.26 - 1.28] | 0.73 [0.73 - 0.74] | 0.84 [0.83 - 0.84] | 0.83 [0.82 - 0.84] |
|  | Jewish | 1.08 [1.06 - 1.10] | 0.99 [0.98 - 1.01] | 1.04 [1.03 - 1.06] | 0.82 [0.81 - 0.84] | 0.96 [0.94 - 0.98] | 0.94 [0.92 - 0.95] |
|  | Muslim | 1.82 [1.81 - 1.83] | 1.63 [1.63 - 1.64] | 1.58 [1.57 - 1.59] | 0.57 [0.56 - 0.57] | 0.61 [0.60 - 0.61] | 0.63 [0.63 - 0.64] |
|  | Sikh | 1.87 [1.85 - 1.89] | 1.74 [1.72 - 1.76] | 1.74 [1.72 - 1.76] | 0.79 [0.78 - 0.80] | 0.85 [0.84 - 0.86] | 0.83 [0.82 - 0.84] |
|  | No religion | 0.84 [0.84 - 0.84] | 0.86 [0.86 - 0.86] | 0.87 [0.87 - 0.87] | 0.95 [0.95 - 0.95] | 0.95 [0.95 - 0.95] | 0.97 [0.97 - 0.97] |
|  | Other religion | 0.77 [0.76 - 0.79] | 0.79 [0.77 - 0.81] | 0.80 [0.78 - 0.82] | 0.75 [0.74 - 0.77] | 0.78 [0.76 - 0.79] | 0.79 [0.77 - 0.80] |
|  | Not stated | 0.87 [0.86 - 0.87] | 0.88 [0.87 - 0.88] | 0.88 [0.87 - 0.89] | 0.86 [0.85 - 0.86] | 0.87 [0.87 - 0.87] | 0.88 [0.87 - 0.88] |
| Household tenure | Owned | 1 (ref) | 1 (ref) | 1 (ref) | 1 (ref) | 1 (ref) | 1 (ref) |
|  | Other | 0.96 [0.95 - 0.97] | 0.97 [0.96 - 0.98] | 0.94 [0.93 - 0.95] | 0.84 [0.83 - 0.84] | 0.87 [0.86 - 0.88] | 0.90 [0.89 - 0.91] |
|  | Private rented | 0.92 [0.92 - 0.93] | 0.91 [0.91 - 0.92] | 0.90 [0.90 - 0.90] | 0.82 [0.82 - 0.82] | 0.84 [0.84 - 0.85] | 0.87 [0.87 - 0.88] |
|  | Social rented | 1.07 [1.07 - 1.08] | 1.02 [1.02 - 1.02] | 0.96 [0.95 - 0.96] | 0.82 [0.82 - 0.82] | 0.84 [0.84 - 0.85] | 0.87 [0.87 - 0.87] |
| Care home status | No | 1 (ref) | 1 (ref) | 1 (ref) | 1 (ref) | 1 (ref) | 1 (ref) |
|  | Yes | 4.77 [4.71 - 4.83] | 4.83 [4.77 - 4.89] | 5.01 [4.95 - 5.08] | 1.04 [1.02 - 1.07] | 1.04 [1.01 - 1.06] | 1.31 [1.27 - 1.35] |
| National Statistics Socio-Economic Classification of the household reference person | 1 Higher managerial, administrative and professional occupations | 1 (ref) | 1 (ref) | 1 (ref) | 1 (ref) | 1 (ref) | 1 (ref) |
|  | 2 Lower managerial, administrative and professional occupations | 1.28 [1.27 - 1.28] | 1.27 [1.26 - 1.28] | 1.23 [1.22 - 1.24] | 1.05 [1.05 - 1.06] | 1.04 [1.03 - 1.04] | 1.03 [1.02 - 1.04] |
|  | 3 Intermediate occupations | 1.37 [1.36 - 1.37] | 1.34 [1.33 - 1.35] | 1.22 [1.21 - 1.23] | 1.01 [1.00 - 1.02] | 0.98 [0.98 - 0.99] | 0.98 [0.98 - 0.99] |
|  | 4 Small employers and own account workers | 1.33 [1.32 - 1.34] | 1.35 [1.34 - 1.36] | 1.20 [1.19 - 1.21] | 0.89 [0.88 - 0.89] | 0.88 [0.87 - 0.88] | 0.90 [0.89 - 0.90] |
|  | 5 Lower supervisory and technical occupations | 1.52 [1.51 - 1.53] | 1.50 [1.49 - 1.51] | 1.33 [1.32 - 1.34] | 0.99 [0.98 - 0.99] | 0.94 [0.93 - 0.95] | 0.95 [0.94 - 0.96] |
|  | 6 Semi-routine occupations | 1.60 [1.59 - 1.61] | 1.56 [1.55 - 1.57] | 1.36 [1.35 - 1.37] | 0.95 [0.94 - 0.95] | 0.90 [0.90 - 0.91] | 0.93 [0.93 - 0.94] |
|  | 7 Routine occupations | 1.54 [1.53 - 1.55] | 1.48 [1.47 - 1.49] | 1.28 [1.27 - 1.29] | 0.92 [0.91 - 0.93] | 0.87 [0.87 - 0.88] | 0.92 [0.91 - 0.92] |
|  | 8 Never worked and long-term unemployed | 1.52 [1.50 - 1.53] | 1.38 [1.37 - 1.39] | 1.06 [1.05 - 1.07] | 0.68 [0.68 - 0.69] | 0.68 [0.67 - 0.68] | 0.77 [0.77 - 0.78] |
| Country of birth | UK | 1 (ref) | 1 (ref) | 1 (ref) | 1 (ref) | 1 (ref) | 1 (ref) |
|  | Non-UK | 1.23 [1.23 - 1.24] | 1.16 [1.15 - 1.16] | 1.19 [1.18 - 1.19] | 0.68 [0.68 - 0.68] | 0.75 [0.74 - 0.75] | 0.79 [0.78 - 0.79] |
| English language proficiency | Main language | 1 (ref) | 1 (ref) | 1 (ref) | 1 (ref) | 1 (ref) | 1 (ref) |
|  | Well or very well | 1.38 [1.37 - 1.39] | 1.28 [1.28 - 1.29] | 1.32 [1.31 - 1.32] | 0.65 [0.64 - 0.65] | 0.71 [0.71 - 0.72] | 0.76 [0.75 - 0.76] |
|  | Not well or not at all | 1.53 [1.52 - 1.54] | 1.38 [1.37 - 1.39] | 1.33 [1.32 - 1.34] | 0.54 [0.54 - 0.55] | 0.58 [0.58 - 0.59] | 0.65 [0.64 - 0.66] |
| Rural-Urban Classification | Villages, hamlets and isolated dwellings | 1 (ref) | 1 (ref) | 1 (ref) | 1 (ref) | 1 (ref) | 1 (ref) |
|  | City and town | 1.40 [1.39 - 1.41] | 1.38 [1.37 - 1.39] | 1.30 [1.30 - 1.31] | 1.08 [1.07 - 1.08] | 1.08 [1.07 - 1.08] | 1.13 [1.13 - 1.14] |
|  | Major or minor conurbation | 1.87 [1.86 - 1.88] | 1.65 [1.64 - 1.66] | 1.49 [1.48 - 1.50] | 0.94 [0.94 - 0.95] | 1.02 [1.02 - 1.03] | 1.12 [1.12 - 1.13] |
|  | Town and fringe | 1.19 [1.18 - 1.20] | 1.16 [1.15 - 1.17] | 1.16 [1.15 - 1.16] | 1.10 [1.09 - 1.11] | 1.09 [1.08 - 1.09] | 1.09 [1.08 - 1.09] |

OR, odds ratio; CI, confidence interval (95%).
Model 1, adjusted for age and sex only; Model 2, plus geography (region and Rural-Urban Classification); Model 3, fully-adjusted model.

For religious affiliation, the highest OR of testing positive for SARS-CoV-2 (relative to the Christian group) was observed for people identifying as Muslim in the second wave; when adjusting for age and sex, the OR was 1.82 (1.81 to 1.83), reducing to 1.58 (1.57 to 1.59) in the fully-adjusted model. This suggests that geography, socio-demographic factors and pre-pandemic health status explained 29.3% of the elevated odds of testing positive for SARS-CoV-2 among people identifying as Muslim during the second wave of the pandemic. During the third wave, the odds of testing positive for SARS-CoV-2 were highest among those identifying as Christian; the lowest OR was observed in the Muslim population at 0.63 (0.63 to 0.64), whilst the highest was for the “No religion” group at 0.97 (0.97 to 0.97).

There were large differences and variations in risk over time according to care home residency status. In the second wave, the fully-adjusted OR of testing positive for people living in a care home was 5.01 (4.95 to 5.08) compared to those not in a care home, whereas in the third wave the fully-adjusted OR was 1.31 (1.27 to 1.35).

Several other factors were also independently associated with SARS-CoV-2 infection. For example, people living in urban areas had higher odds of testing positive for SARS-CoV-2 than those living in rural areas during both the second and third waves. Living in a more deprived area was also associated with a higher odds of testing positive during the second wave but not in the third wave. During the second wave, people who reported that English was not their main language had higher odds of testing positive for SARS-CoV-2 than those who reported speaking English as their main language after adjusting for confounding factors. Conversely, during the third wave, the odds of testing positive among people who did not speak English as their main language was lower than for native English speakers. Disabled people who were limited a lot in their daily activities had elevated odds of testing positive during the second wave after adjusting for age and sex only but had lower odds than non-disabled people in the fully-adjusted model. In the third wave, disabled people had lower odds of testing positive than non-disabled people across all models.

After stratifying the analyses by broad age group (under 65 years versus 65 years and over), larger associations were observed among people aged 65 and over for many socio-demographic characteristics than among those aged under 65 (Tables 4 and 5). Notably, among people aged under 65 years, all ethnic minority groups had lower odds of testing positive than the White British group during the third wave. Conversely, during the third wave among people aged 65 years and over, the fully-adjusted odds of testing positive were notably higher for the Bangladeshi (OR 1.63, 1.52 to 1.75) and Pakistani (1.35, 1.30 to 1.40) ethnic groups than for the White British. A similar pattern was observed for religious affiliation; during the third wave, the odds of testing positive were lower than the Christian group for all other religions among people aged under 65 years but were higher among people aged 65 years and over in the fully-adjusted models for people identifying as Sikh, Hindu, Muslim, or Jewish. Disabled people aged under than 65 years had lower odds of testing positive than non-disabled people in both the second and third waves, whereas disabled people aged 65 years and over had higher odds than non-disabled people in the second wave but were not at elevated risk of testing positive in the third wave after adjustment.

**Table 4:**
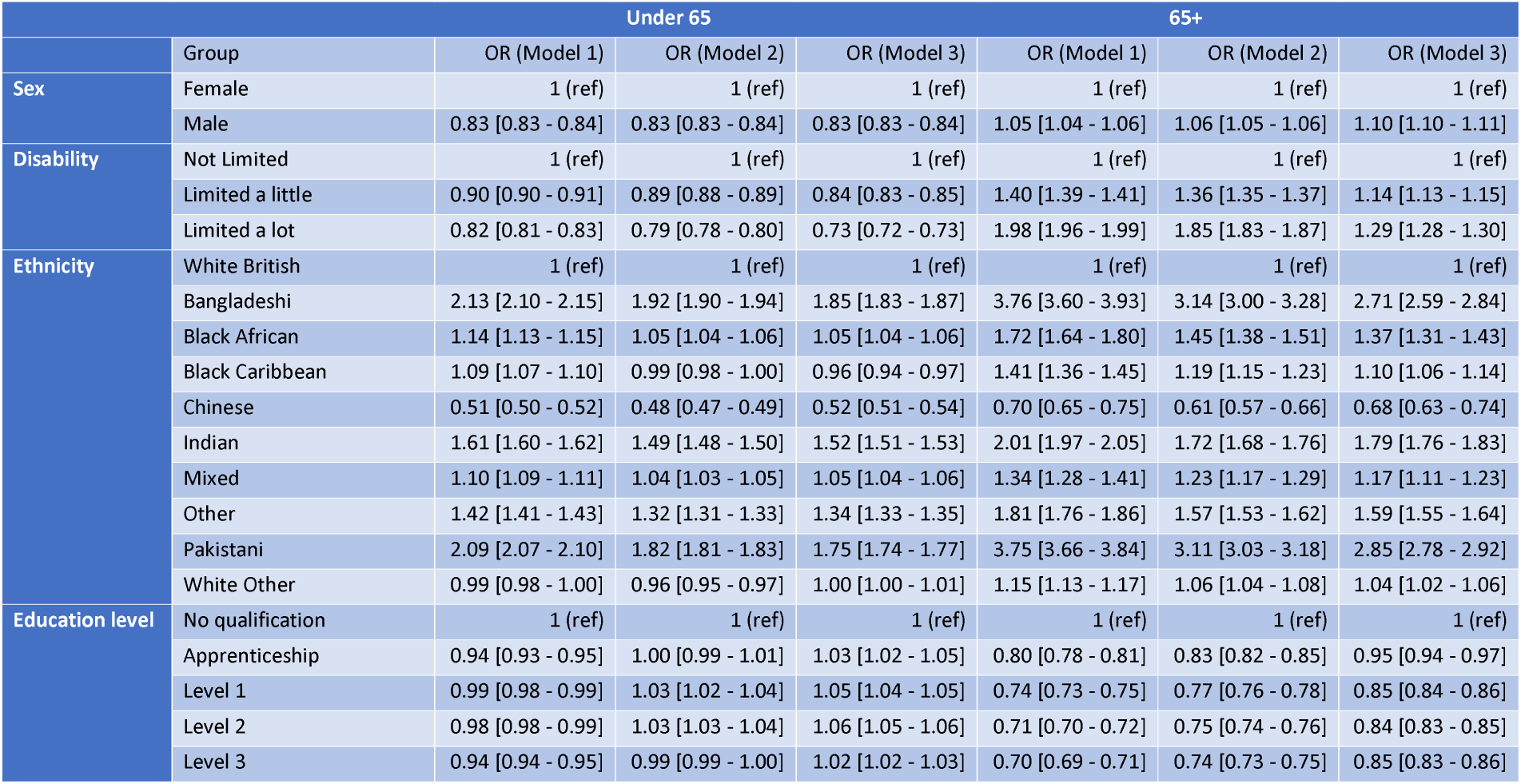
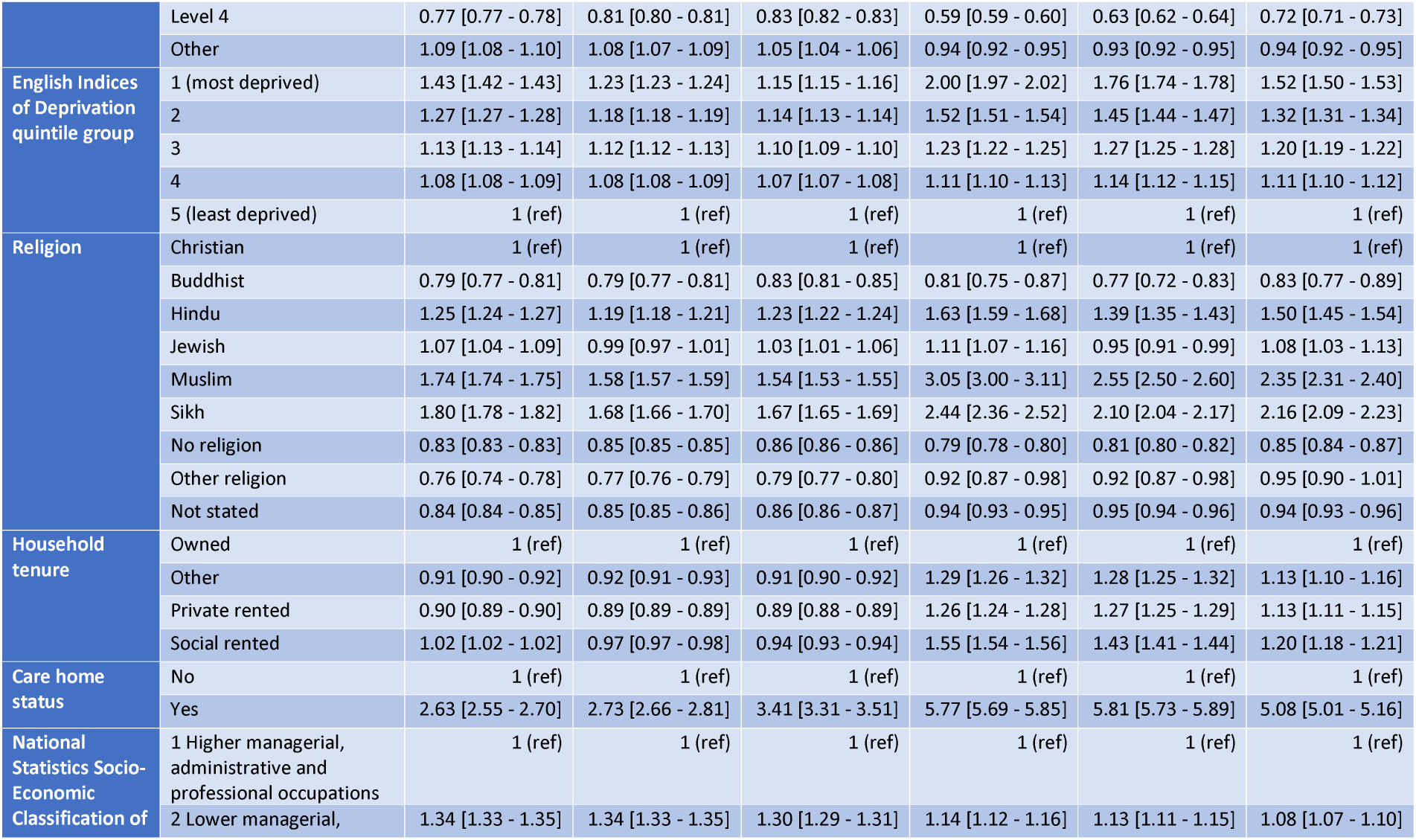
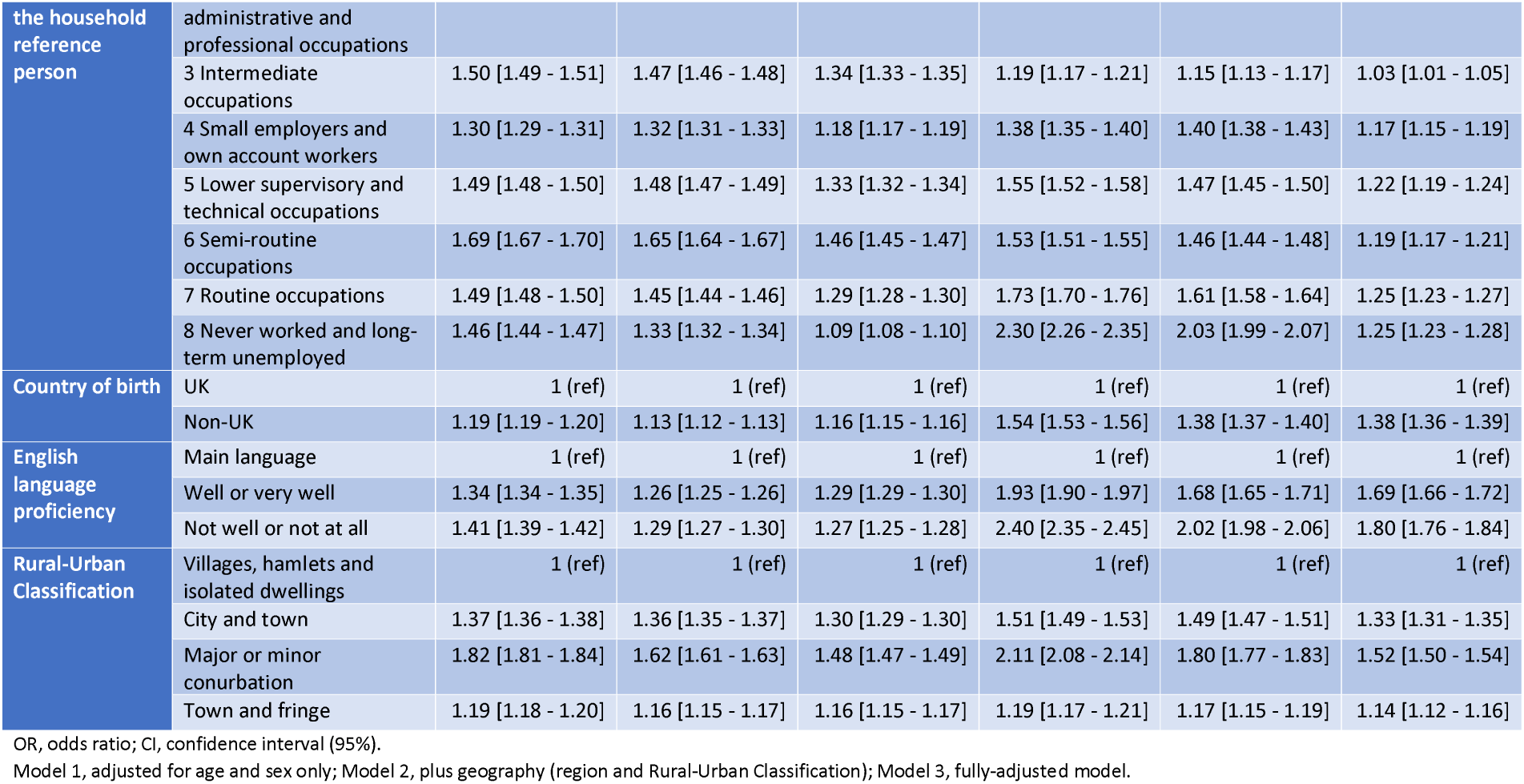
Adjusted odds ratios of receiving a positive test for SARS-CoV-2 by sociodemographic characteristics and age group during the second wave (1 September 2020 to 22 May 2021)

**Table 5:**
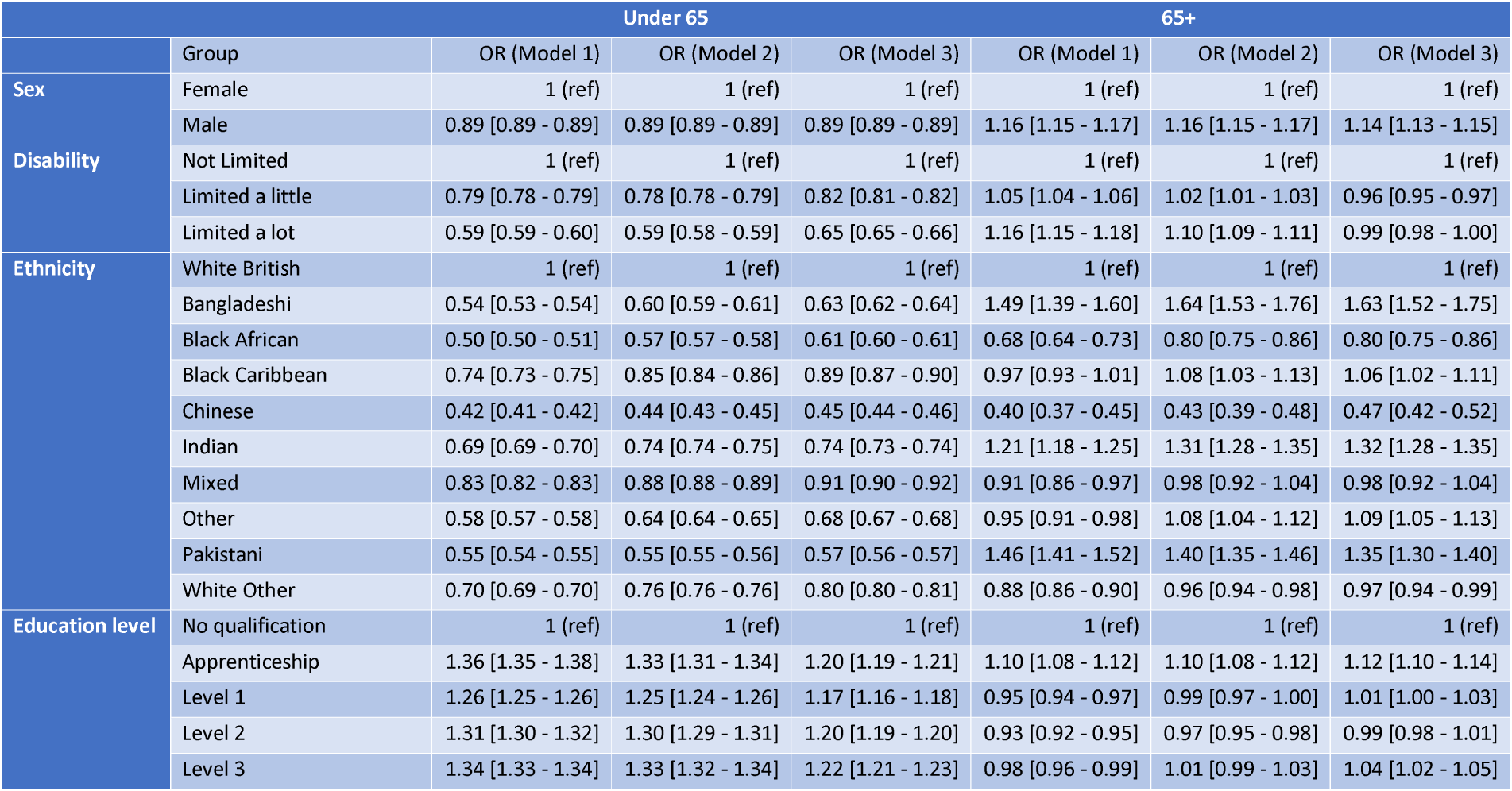
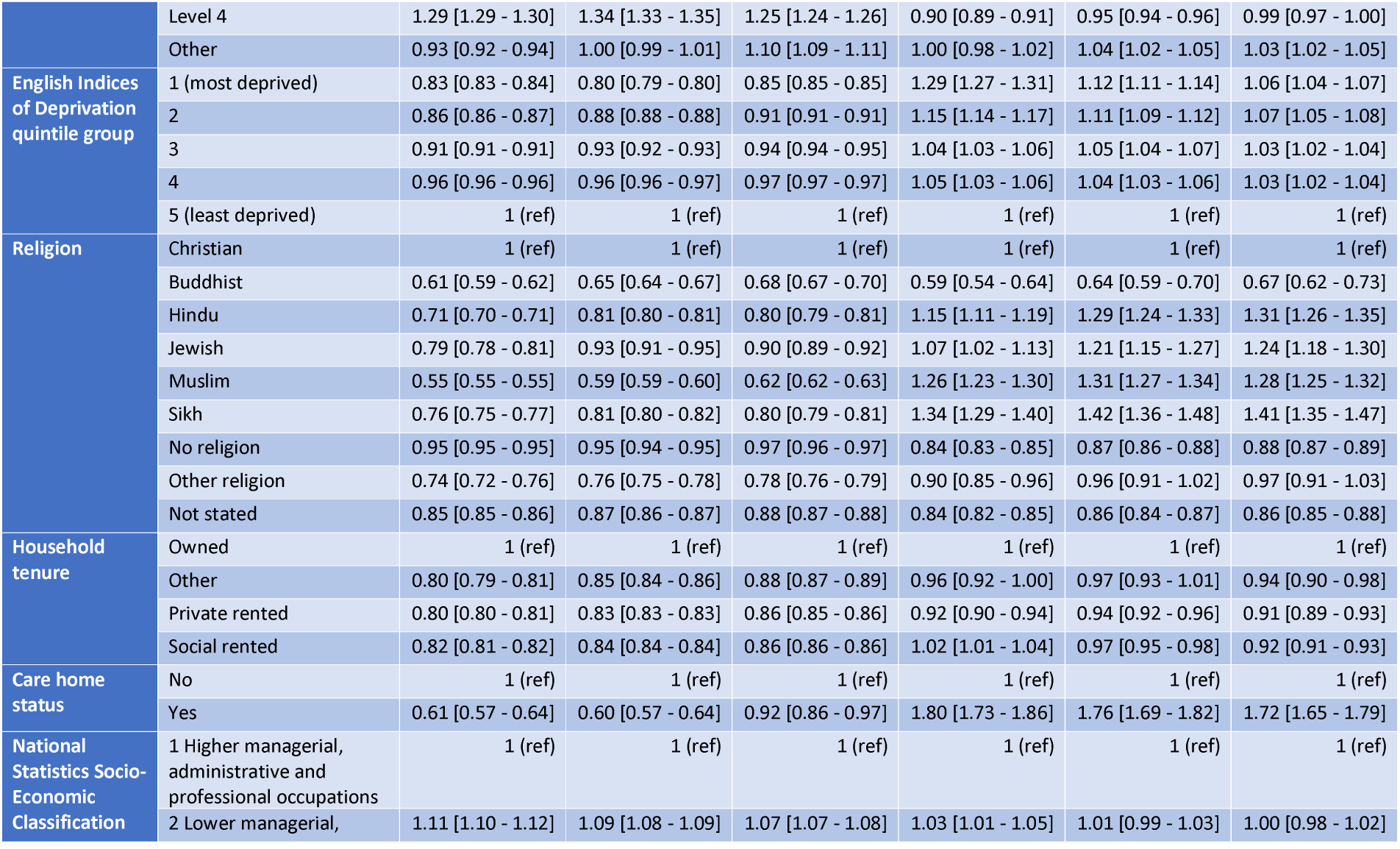
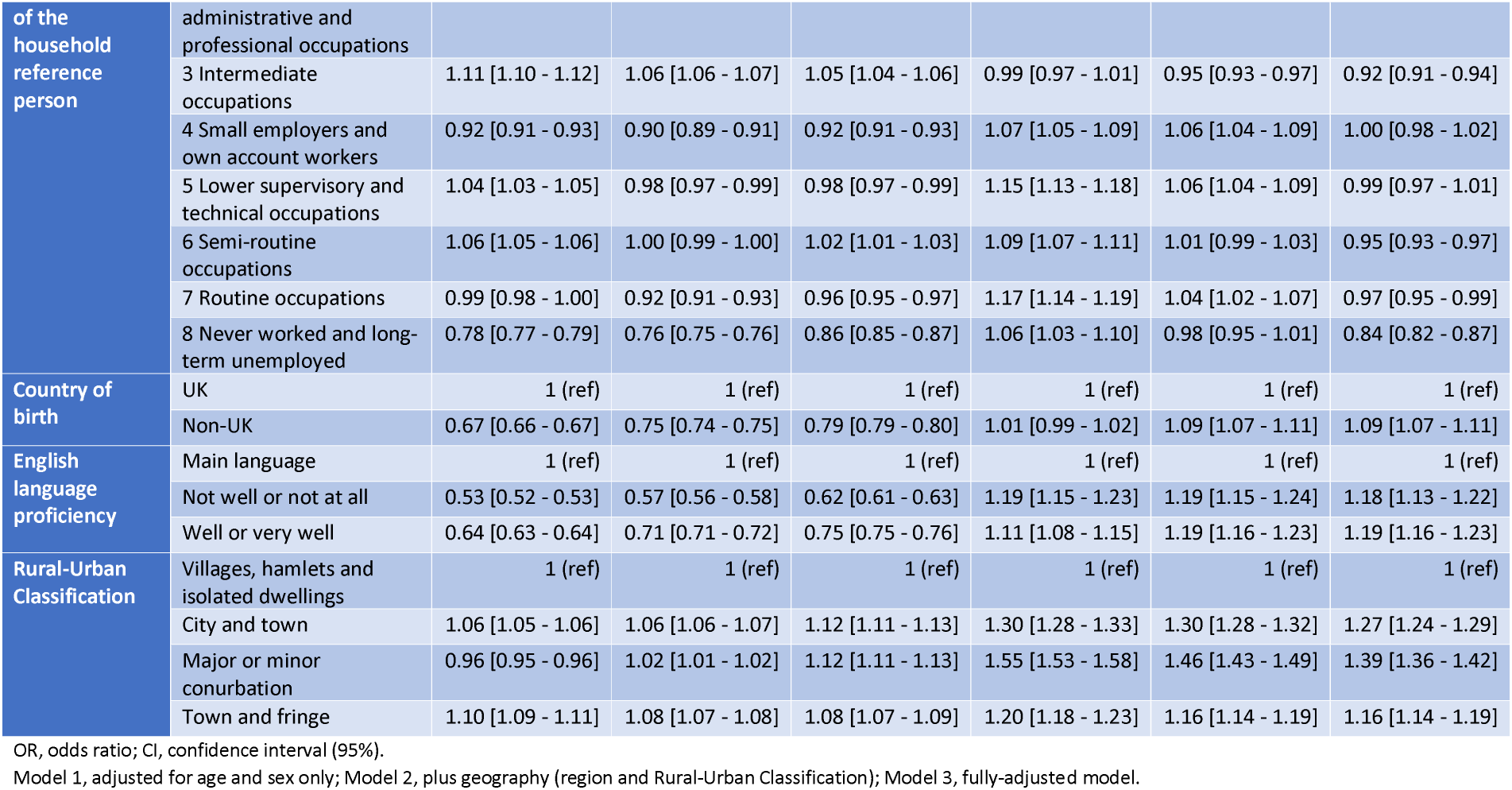
Adjusted odds ratios of receiving a positive test for SARS-CoV-2 by sociodemographic characteristics and age group during the third wave (23 May 2021 to 10 December 2021)

## Discussion

### Main findings

Our analysis using population-level linked data in England shows that there were significant disparities in COVID-19 case rates in people aged ≥10 years during the Alpha and Delta waves for several socio-demographic characteristics, most notably by ethnic group, religious affiliation, and Rural-Urban Classification. During the second wave, case rates were highest among Bangladeshi and Pakistani ethnic groups, with adjustments for geography, socio-economic factors and pre-existing health conditions accounting for 26% and 31% of the excess risk, respectively. For religious affiliation, those who identified as Muslim or Sikh had the highest rates, with adjustments only accounting for 27% and 17% of the excess risk, respectively. While some differences were found by deprivation and other socio-demographic factors, these were less pronounced than for ethnicity or religious affiliation. However, there is significant overlap between ethnicity and religion; 93.4% of people from both the Pakistani and Bangladeshi ethnic groups within the study self-identified as Muslim.

For the third wave – corresponding to the emergence of the Delta variant – we observed a different pattern for several factors. The White British ethnic group had the highest case rates and odds ratios, whilst those who self-identified as Christian had the highest rates among religious affiliations. Rates also became highest among people born within the UK and whose main language was English, with cases particularly prevalent among younger age groups. A potential reason is that levels of population immunity were higher for the groups that had the highest case rates in the first and second waves. However, when stratifying these results by broad age groups (under 65 years versus 65 years and over), we found that the odds for all ethnic minority groups were higher among the elderly. This could in part be due to ethnic minority groups, particularly those of Bangladeshi, Pakistani or Indian ethnic background, being more likely to live in multigenerational and larger households, which may partly explain the continued elevated risk of mortality during the third wave for ethnic minority groups relative to the White British population [12, 30].

### Comparison with other studies

Our findings are consistent with results from the Coronavirus Infection Survey, which found that between September 2020 to May 2021, individuals living in urban areas and deprived areas, and of a younger age were most likely to test positive in the UK [16]. Studies using UK COVID-19 surveillance data have also suggested that Black and South Asian ethnic groups were more likely to test positive than White British individuals in England [6] [31]. In addition, our results support previous analyses using UK administrative data that have shown higher age-standardised case rates among ethnic minority groups until June 2021, when rates increased among the White population [30]. Similar patterns of increased infection in the most deprived areas and among minority ethnic groups have been observed worldwide [32] [10].

Studies have shown that COVID-19 vaccinations significantly reduced the risk of SARS-CoV-2 infection [18]. In addition, from December 2020 onwards, unadjusted vaccination uptake rates have been lower among adults from ethnic minority groups, those living in the most deprived areas, self-reporting being disabled, of younger age, did not speak English as their first language, and belonged to a lower socio-economic group [30] [33]. This is consistent with our findings when adjusting for age and sex only during the second wave, suggesting that lower vaccine uptake rates for certain groups and younger people might contribute to case rate inequalities. Although vaccination rates were lower for the Bangladeshi and Pakistani groups than the White British population, the lowest rates were found in Black African and Black Caribbean groups.

### Strengths and limitations

The primary strength of the study is using nationwide linked population-level data that combines a diverse set of demographic and socio-economic factors from the 2011 Census with timely data on national SARS-CoV-2 testing. Unlike studies based solely on electronic health records, our study is based on self-identified ethnicity, limiting the potential for exposure misclassification bias. We also have information on a wide range of socio-demographic factors not typically available in electronic health records, such as religion, main language and educational attainment. Another strength is the size of the dataset, comprising 78.4% of people aged 10 years and over living in England in 2020. Therefore, this study is sufficiently powered to detect small differences in the odds of testing positive for SARS-CoV-2 by detailed characteristics after adjusting for confounding factors and interactions with age.

An important limitation is that the PHDA only contains information on people who were enumerated at the 2011 Census. It therefore excludes people living in England in 2011 but who did not participate in the 2011 Census (estimated to be approximately 5% of the population at the time); respondents who could not be linked to the 2011 to 2013 NHS Patient Registers (5.4% of Census respondents); people who have immigrated since 2011; children younger than 10-years-old in 2021; and people not registered with a GP surgery or who had opted out of GDPPR.

A further limitation is that many of the socio-demographic variables were derived from the 2011 Census. Some of these characteristics (for example disability status and NS-SEC) might have changed since the 2011 Census and may not accurately reflect individuals’ circumstances during the pandemic. A possible contributing factor for the inequalities in case rates by ethnicity which is unaccounted for in our study could be the size of the household, with Pakistani and Bangladeshi groups being most likely to live in larger households [34].

National SARS-CoV-2 testing data do not provide a representative measure of infections because people are more likely to get a test for COVID-19 if they have symptoms, as they are advised to do so, and because there may also be other biases in the choice to get a test. Approximately 40% of people who test positive in the Coronavirus Infection Survey do not develop symptoms within 35 days of testing positive [28]. Therefore, these figures are likely to under-represent the number of asymptomatic cases and so may not be generalisable to all infections in the population. In addition, people in certain occupations and school children are required to undergo regular testing, so may be more likely to test positive for COVID-19 as a result of higher testing rates. Adherence to testing has been shown to be lower among males, those of younger age, and people of lower socioeconomic status [35], meaning disparities in case rates are likely to be underestimated.

## Conclusion

There are significant differences in SARS-CoV-2 case rates by various socio-demographics, particularly ethnicity and religion, between different waves of the pandemic. Further research is now needed to understand why these disparities exist and how they can best be addressed through policy interventions. Continued surveillance is essential to ensure that changes in the patterns of infection are identified early to inform public health interventions.

## Supporting information

Supplementary tables and plots

## Data Availability

Information on data availability and access is available via the Secure Research Service: https://www.ons.gov.uk/aboutus/whatwedo/statistics/requestingstatistics/approvedresearcherscheme

