## Supplementary tables and plots for "Disparities in SARS-CoV-2 case rates by ethnicity, religion, measures of socio-economic position, English proficiency, and self-reported disability: cohort study of 39 million people in England during the Alpha and Delta waves"

### Supplementary material

Table S1: Sources of variables used in the analyses

| Variable | Coding | Source |
| --- | --- | --- |
| Positive test for SARS-CoV-2 | Tested positive for SARS-CoV-2 between 1 September 2020 and 10 December 2021 and result recorded in national testing data | NPEx/SGSS |
| Age | Restricted natural cubic spline (using 10-year age bands) | 2011 Census |
| Sex | Female, male | 2011 Census |
| Ethnicity | White British, Bangladeshi, Black African, Black Caribbean, Chinese, Indian, Mixed, Other, Pakistani, White Other | 2011 Census |
| Religious affiliation | Christian, Buddhist, Hindu, Jewish, Muslim, Sikh, no religion, other religion, religion not stated | 2011 Census |
| Region | North East, North West, Yorkshire and The Humber, East Midlands, West Midlands, East of England, London, South East, South West | GDPPR |
| Rural-Urban Classification | Major or minor conurbation; city and town; town and Fringe; villages, hamlets and isolated dwellings | GDPPR |
| English Indices of Deprivation | Dummy variables representing quintile groups of deprivation | GDPPR |
| Residence type | Dummy variables representing private household or other communal establishment, or care home residency | GDPPR/2011 Census |
| Household tenure | Own, social rented, private rented, other | 2011 Census |
| Country of birth | UK, non-UK | 2011 Census |
| English language proficiency | Main language, other | 2011 Census |
| Level of highest qualification | Degree, A-level or equivalent, GCSE or equivalent, no qualification, other | 2011 Census |
| Disability | Non-disabled, disabled (daily activities limited a little), disabled (daily activities limited a lot) | 2011 Census |
| Body Mass Index (kg/m2) | < 18.5, 18.5 to <25, 25 to <30, >= 30, unknown | GDPPR |
| Learning disability | No learning disability, Down’s syndrome, other learning disability | GDPPR |
| Pre-existing conditions | Number of pre-existing conditions | GDPPR/HES |

NPEx, National Pathology Exchange; SGSS, Second Generation Surveillance System; GCSE, General Certificate of Secondary Education; GP, general practitioner; GDPPR, General Practice Extraction Service Data for Pandemic Planning and Research; HES, Hospital Episode Statistics; NHS, National Health Service.

Plot S1: Odds ratios by wave of the pandemic – age and sex

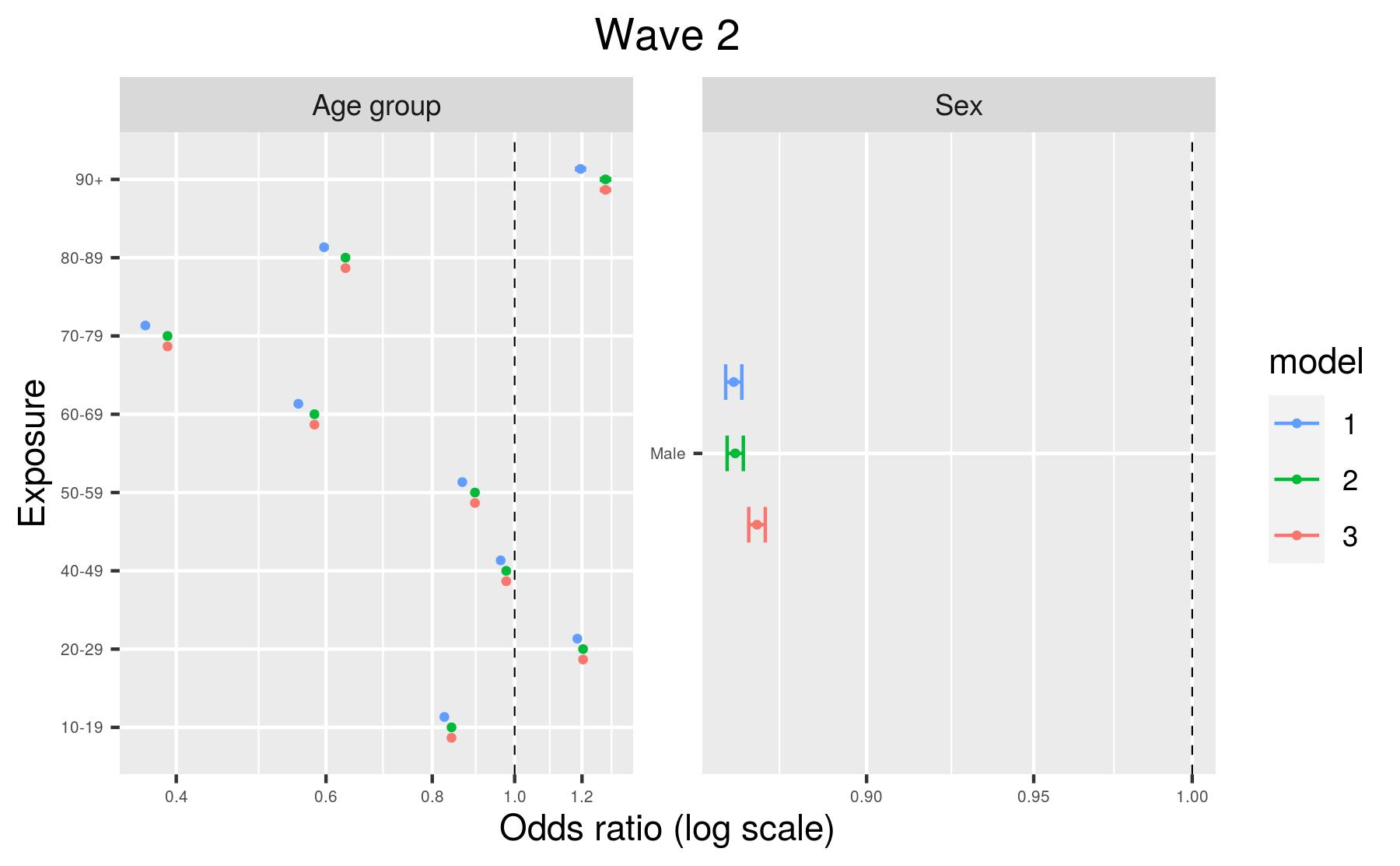

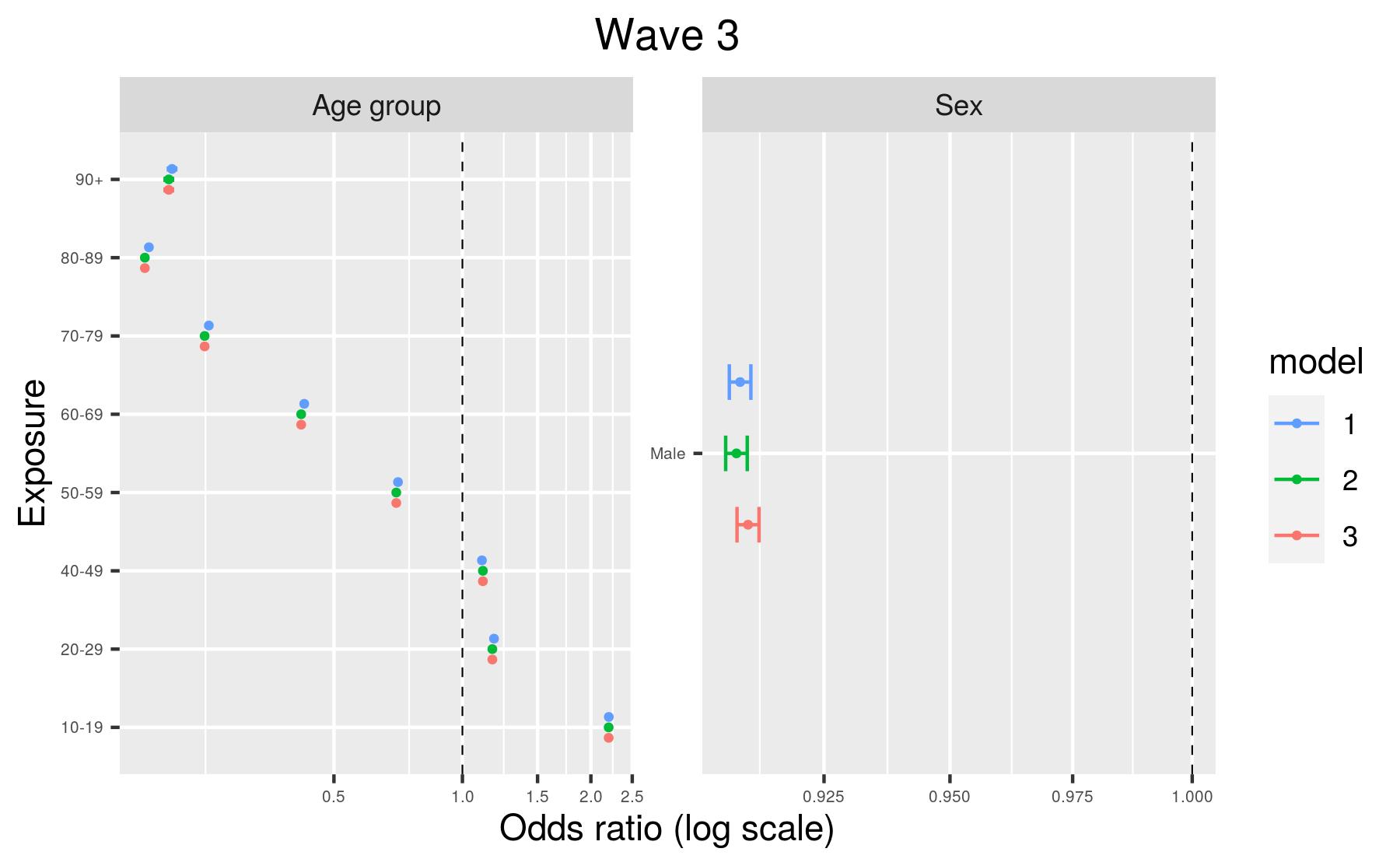

Model 1, adjusted for age and sex only; Model 2, plus geography (region and Rural-Urban Classification); Model 3, fully-adjusted model.

Plot S2: Odds ratios by wave of the pandemic – Geographical variables
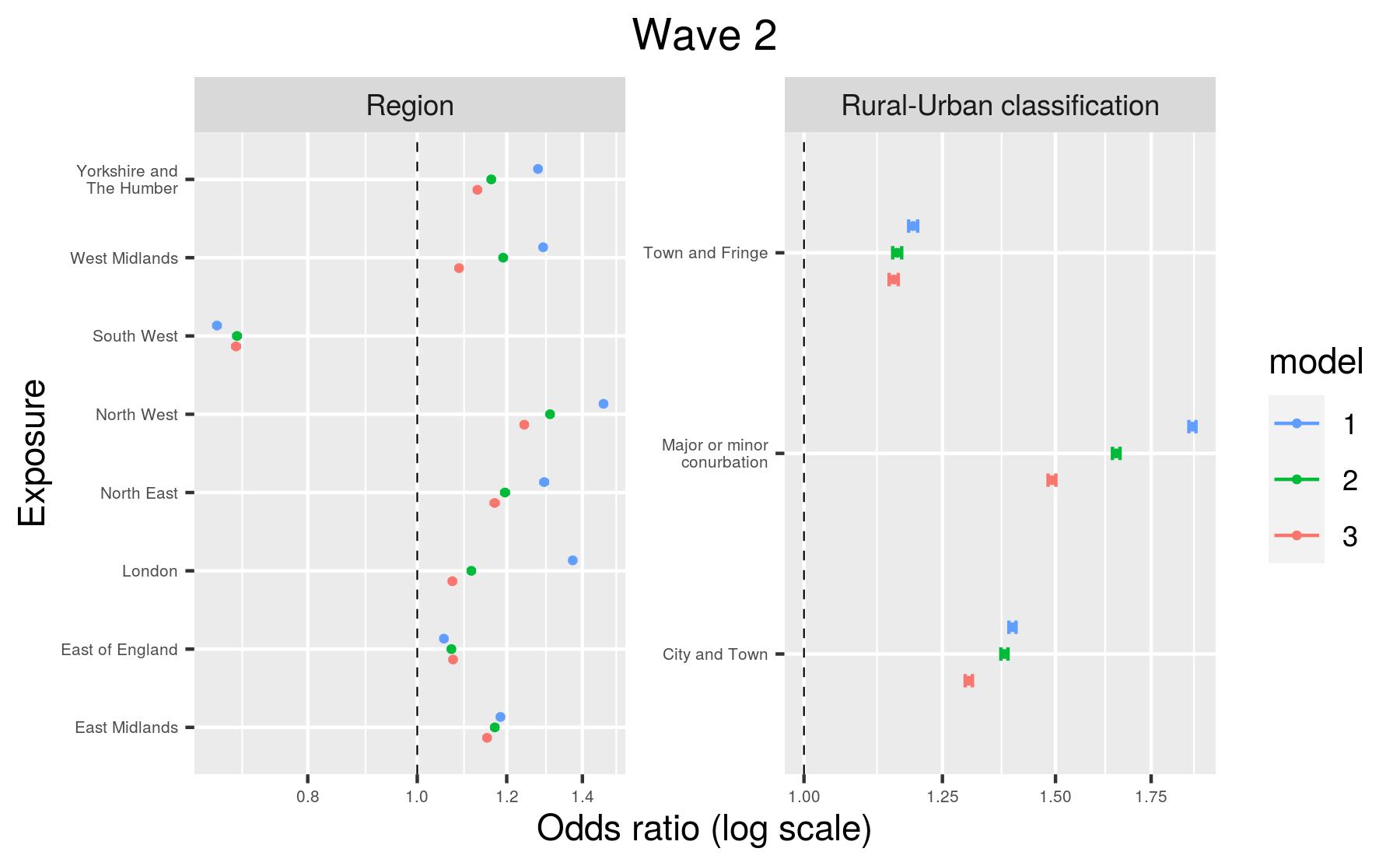

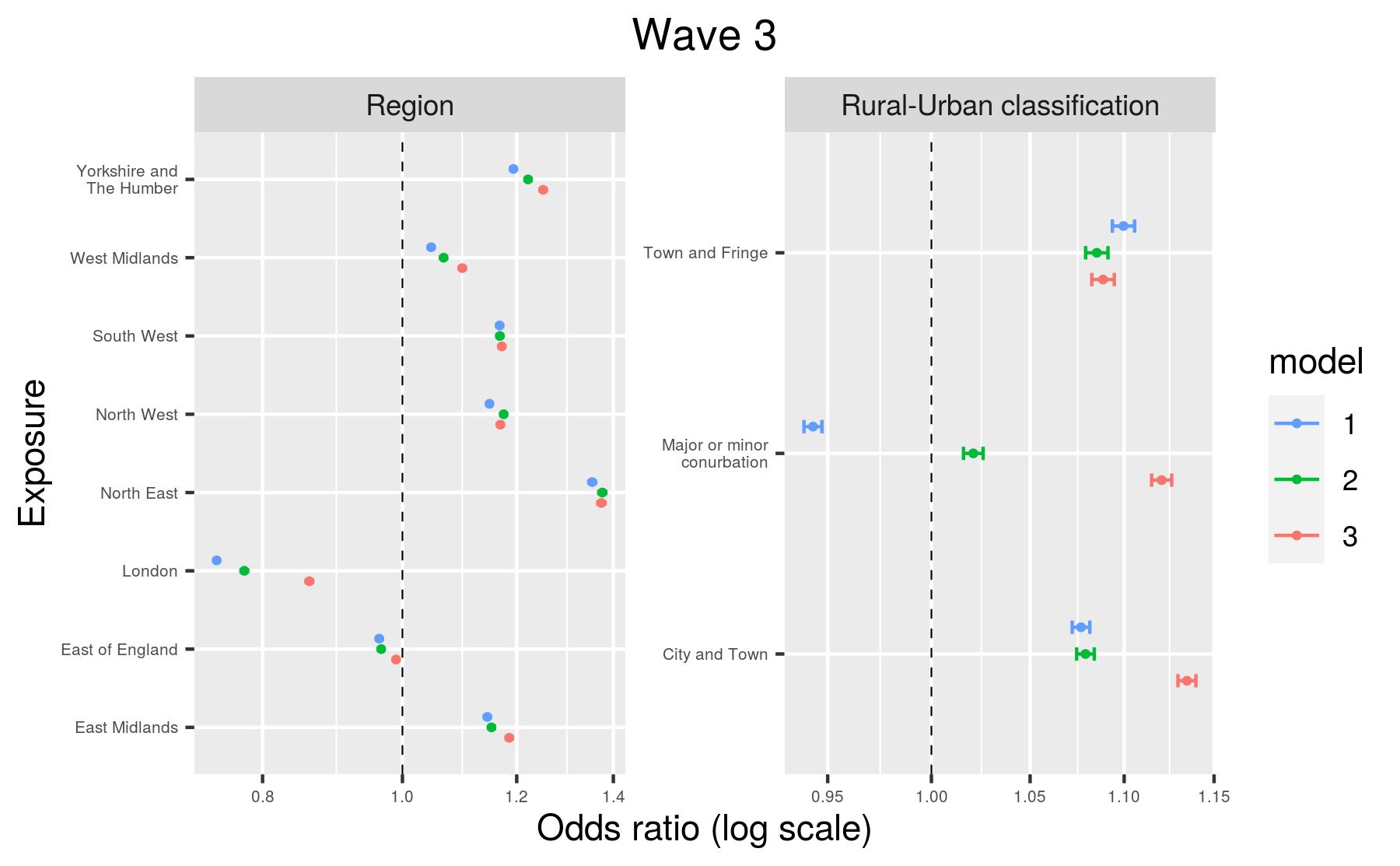

Model 1, adjusted for age and sex only; Model 2, plus geography (region and Rural-Urban Classification); Model 3, fully-adjusted model.

Plot S3: Odds ratios by wave of the pandemic – sociodemographic variables
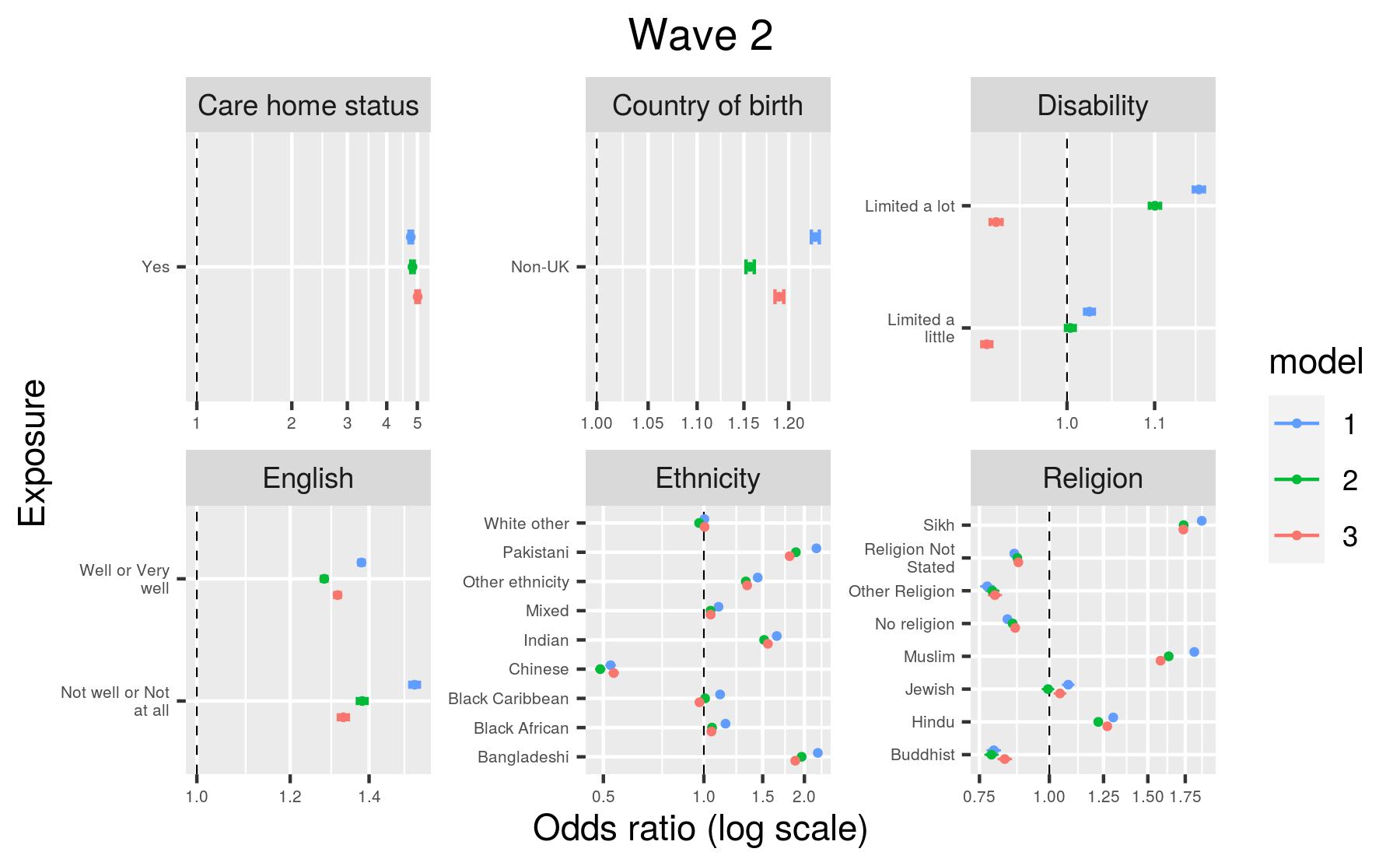

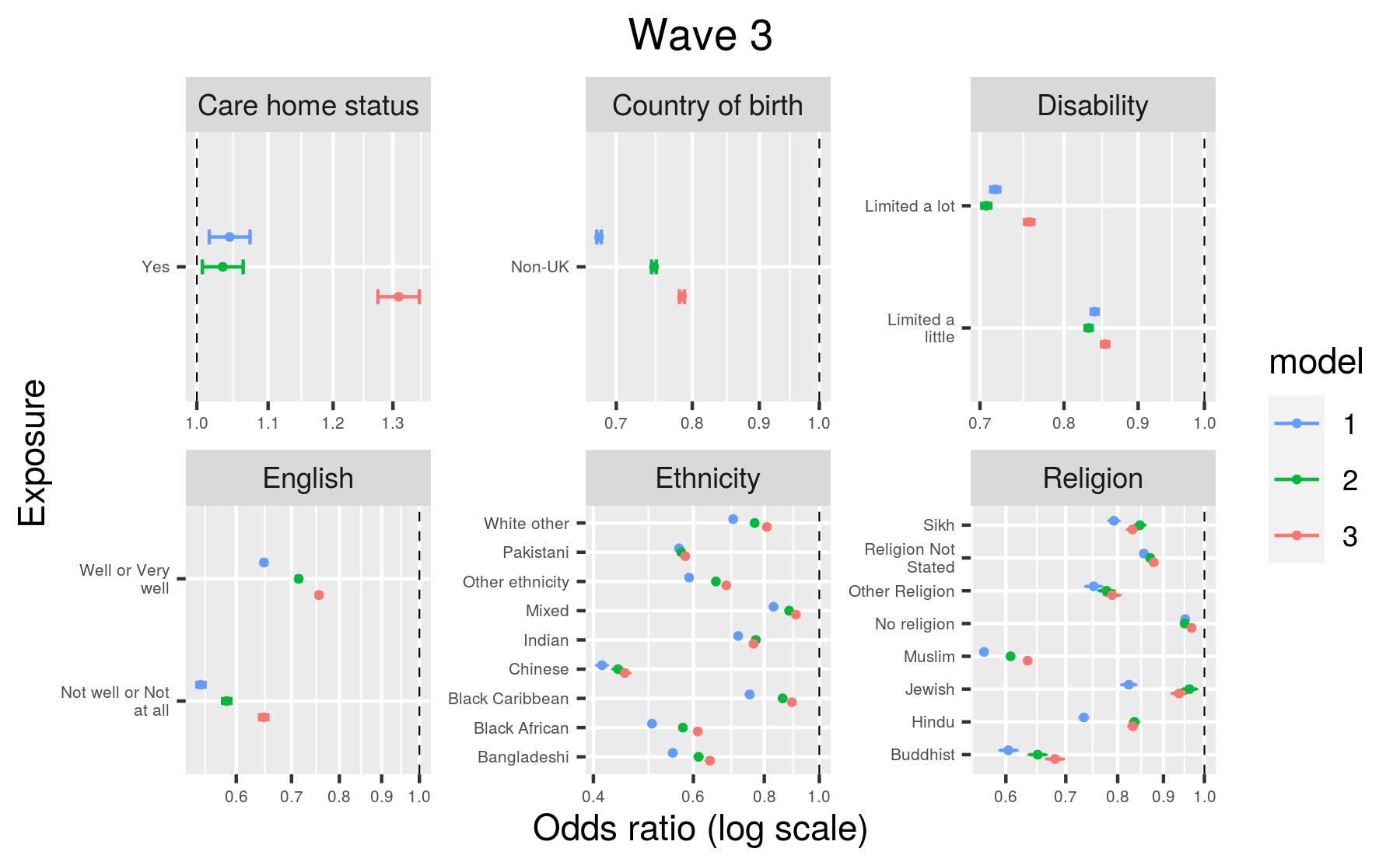

Model 1, adjusted for age and sex only; Model 2, plus geography (region and Rural-Urban Classification); Model 3, fully-adjusted model.

Plot S4: Odds ratios by wave of the pandemic – socioeconomic variables
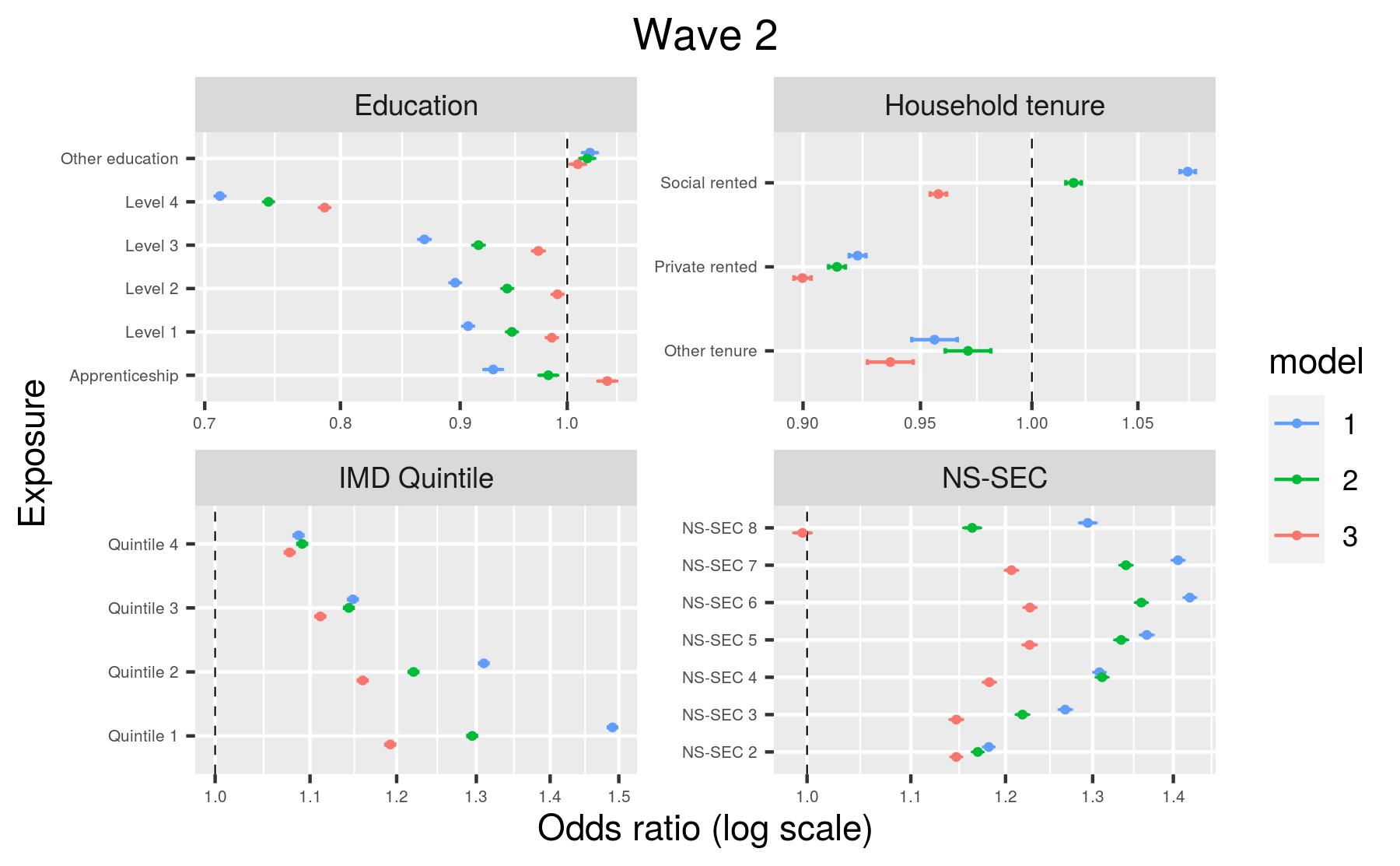

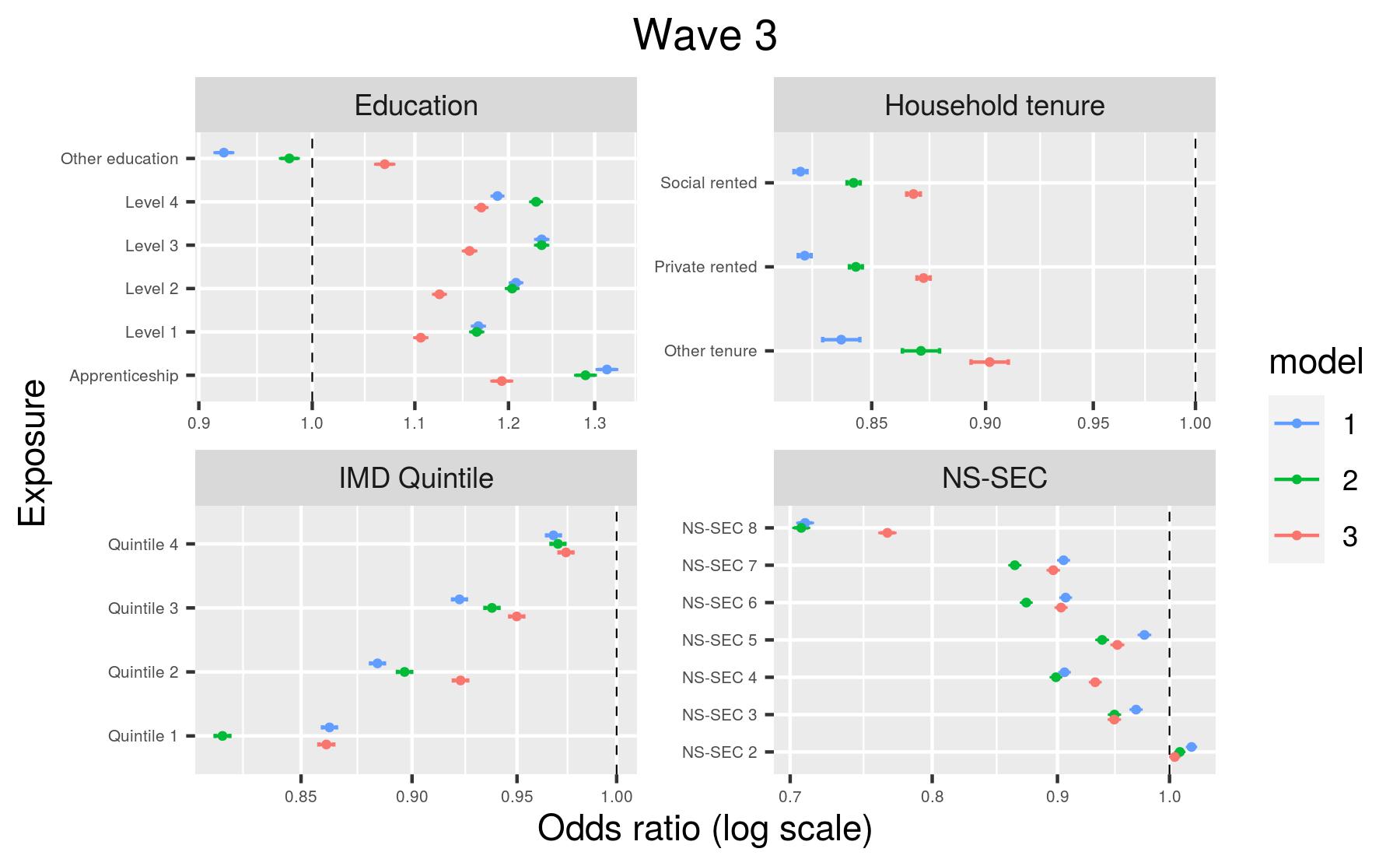

Model 1, adjusted for age and sex only; Model 2, plus geography (region and Rural-Urban Classification); Model 3, fully-adjusted model.
